# Connecting genomic results for psychiatric disorders to human brain cell types and regions reveals convergence with functional connectivity

**DOI:** 10.1101/2024.01.18.24301478

**Authors:** Shuyang Yao, Arvid Harder, Fahimeh Darki, Yu-Wei Chang, Ang Li, Kasra Nikouei, Giovanni Volpe, Johan N Lundström, Jian Zeng, Naomi Wray, Yi Lu, Patrick F Sullivan, Jens Hjerling-Leffler

## Abstract

Understanding the temporal and spatial brain locations etiological for psychiatric disorders is essential for targeted neurobiological research. Integration of genomic insights from genome-wide association studies with single-cell transcriptomics is a powerful approach although past efforts have necessarily relied on mouse atlases. Leveraging a comprehensive atlas of the adult human brain, we prioritized cell types via the enrichment of SNP-heritabilities for brain diseases, disorders, and traits, progressing from individual cell types to brain regions. Our findings highlight specific neuronal clusters significantly enriched for the SNP-heritabilities for schizophrenia, bipolar disorder, and major depressive disorder along with intelligence, education, and neuroticism. Extrapolation of cell-type results to brain regions reveals important patterns for schizophrenia with distinct subregions in the hippocampus and amygdala exhibiting the highest significance. Cerebral cortical regions display similar enrichments despite the known prefrontal dysfunction in those with schizophrenia highlighting the importance of subcortical connectivity. Using functional MRI connectivity from cases with schizophrenia and neurotypical controls, we identified brain networks that distinguished cases from controls that also confirmed involvement of the central and lateral amygdala, hippocampal body, and prefrontal cortex. Our findings underscore the value of single-cell transcriptomics in decoding the polygenicity of psychiatric disorders and offer a promising convergence of genomic, transcriptomic, and brain imaging modalities toward common biological targets.

## Introduction

Genome-wide association studies (GWAS) have yielded fundamental insights into the nature of a wide range of human diseases, disorders, biomarkers, and traits. A recent summary ^1^ of 4,593 GWAS publications studying 3,908 phenotypes found 156,556 significant SNP-trait associations; notably, only 4.19% of significant SNPs were in a protein-coding region. GWAS have been particularly informative for psychiatric disorders whose enigmatic nature has long impeded progress. This body of work has shown that major psychiatric disorders are heritable, that clinically dissimilar disorders nonetheless have genetic overlap, and that this can clarify causality ^2–6^.

However, the genetic architectures of psychiatric disorders have proven to be particularly complex ^7^. For example, predictions that genomic studies of schizophrenia would readily identify a few genes with near-causal effects ^8–10^ are inconsistent with the accumulated results: empirical studies of common genetic variation, rare copy number variation, rare exonic variation (both *de novo* and inherited), and whole genome sequencing ^11–14^ were well-powered to detect a few causal genes shared by most cases and yet none were identified. Compared to many other human diseases/disorders, schizophrenia is notably polygenic ^15^ with the major population impact resulting from inheritance of a large number of common variants of small effect ^11,13,16^. Indeed, the most recent GWAS for schizophrenia ^11^ implicated 287 genomic loci (median size 652 kb, interquartile range, IQR, 238-652 kb) often intersecting multiple protein-coding genes (median 2, IQR 1-6, and 13% of all loci contained no protein-coding genes). It is highly likely that there are many more loci to be discovered. It is not clear how these findings inform understanding of the fundamental nature of schizophrenia or what the underlying neurobiology might be.

In this paper, we evaluate the evidence that genomic results for brain disorders, diseases, and traits point at specific brain cell types. We evaluate the overarching hypothesis that cell types – and not a few major genes – are a principal readout of genomic studies for notably complex psychiatric disorders. We ^17,18^ and others ^19–26^ have previously evaluated this idea. However, for brain traits, the key limitations have been the sheer complexity of the brain and limited transcriptomic data that previously forced reliance on mouse brain transcriptomic surveys. Siletti et al. ^27^ recently published the most comprehensive human transcriptomic dataset to date: single-nucleus RNA sequencing (snRNAseq) of 3.369 million nuclei from 106 anatomical dissections within 10 brain regions.

Here, we incorporate the first large-scale human brain atlas along with newer and larger GWAS. We extend our prior work by evaluating evidence for anatomical regions as well as their functional connectivity. Cell types form local networks that connect distributed brain regions. Functional magnetic resonance imaging (fMRI) is a non-invasive and widely used tool to evaluate brain regional functional connectivity in both health and disease. Systematic reviews have suggested disturbances in the Default Mode Network and the Core Network in cases with schizophrenia and their neurotypical relatives ^28–30^. These findings suggest that genetic liability to schizophrenia can be manifest in empirically-defined cell types, anatomical regions, and in functional connectivity between brain regions. In this paper, we integrate fMRI data from schizophrenia cases and controls to illustrate how genetic information, via transcriptomics, agrees with fMRI in the prioritization of brain regions and changes in connectivity. Identifying affected brain regions and circuits is important given “interventional psychiatry” therapeutics that can modulate activity of specific brain regions (e.g., transcranial magnetic stimulation or deep brain stimulation).

## Results

Our overarching goal was to evaluate whether the genomic regions identified by GWAS for complex brain phenotypes implicated specific brain cell types, anatomical regions, or their connectivity. As diagrammed in *Figure 1*, we integrated the most comprehensive human snRNAseq brain atlas to date ^27^ with GWAS summary statistics for 36 primary traits including psychiatric disorders, brain traits, neurological diseases, structural MRI measures, and control traits (*Table S1*, *Figure S1*, and *Methods* for inclusion criteria). We systematically processed summary statistics for these GWAS. *Figure S2* shows the genetic correlations between the primary traits which were in accord with prior reports ^3^. As in our past papers ^17,18^, we used stratified LD score regression (S-LDSC) to estimate the enrichment of SNP-heritability for a trait in genes whose expression typified cell classes. The genetic liability of a trait can be measured by SNP-heritability, the proportion of phenotypic variance in a trait attributable to the additive genetic variation estimated from GWAS data ^31^.

**Figure 1.**
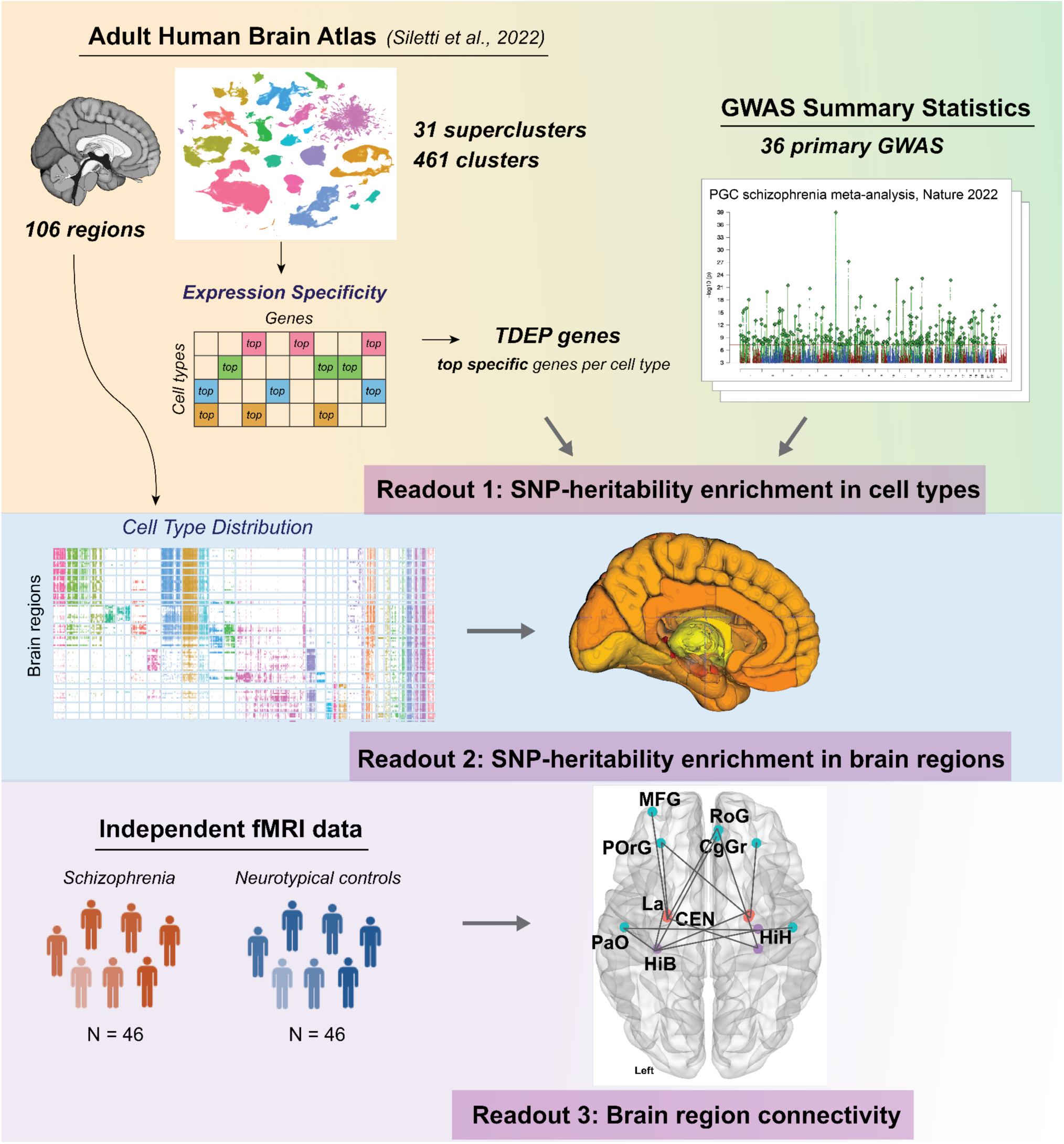
Study schematic. We first identified cell types enriched for the SNP-heritability of 36 primary traits including major psychiatric disorders, using the most comprehensive Adult Human Brain Atlas. This was integrated with the cell type distribution across brain regions to identify brain regions enriched for the SNP-heritability of the traits. Finally, the regions suggested by cell-type-informed SNP-heritability enrichment were used to explore brain region connectivity that can differentiate schizophrenia from neurotypical controls in an independent sample. TDEP=top decile expression proportion, which were the most specifically expressed genes in each cell type (supercluster or cluster).

**Figure 2.**
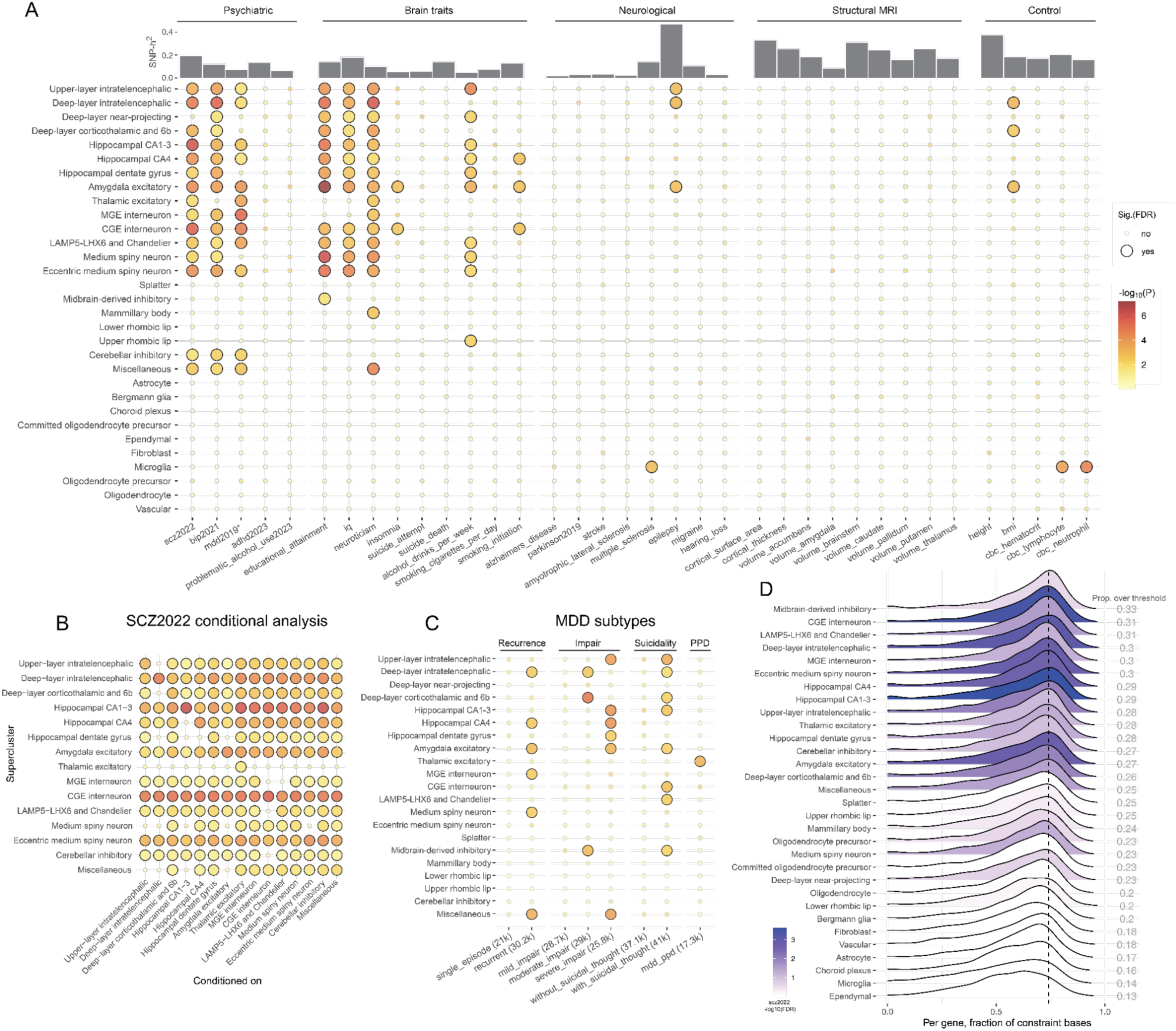
Supercluster results. (A) SNP-heritability enrichment in 31 superclusters for five phenotype categories (psychiatric disorders, brain traits, neurological disorders, structural MRI, and control). Large dots indicate enrichment significance, FDR ≤ 0.05. Dot color indicates enrichment significance as –log10(P) with darker reds indicating greater significance. We interpret the suicide results cautiously as this GWAS did not clearly control for MDD ^40^ which may confound the enrichment pattern. mdd2019* summary statistics did not include data from 23andMe. (B) Conditional analysis of superclusters for enrichment of scz2022 SNP-heritability. The conditional analysis was performed in a pairwise fashion (Methods) for the significant superclusters. The y-axis is the supercluster of interest and the x-axis indicates the supercluster conditioned upon. For convenience, the unconditional results are on the diagonal. Significance was unchanged for deep-layer intratelencephalic, hippocampal CA1-3, amygdala excitatory, CGE interneuron, and eccentric medium spiny neuron, indicating the statistical independence of the results (Table S4). (C) SNP-heritability enrichment for MDD subtypes. Number of cases is shown in parenthesis in the x-axis label. Non-neuronal superclusters did not have signals for any subtype and were therefore omitted in the plot. Dot color and size are the same across panels A-C. (D) Ridge plot showing the density of the evolutionary constraint for TDEP genes of each supercluster. For each gene, the proportion of constraint1 in its CDS bases was used as the measure of evolutionary constraint. The vertical dashed line shows the 80th percentile for evolutionary constraint for all protein-coding genes. The right column gives the proportion of TDEP genes above the 80th percentile of constraint. The plots were colored by the SNP-heritability enrichment for scz2022 (-log_10_FDR).

Cellular diversity is hierarchically organized in the brain ^32,33^, from a tripartite classification (neuronal excitatory, neuronal inhibitory, and non-neuronal) to higher-order cell superclusters that are divisible into clusters and subclusters/cell types. Following the Siletti nomenclature ^27^, we analyzed 31 superclusters and their component 461 clusters. In *Table S2*, we characterize the superclusters: 10 non-neuronal and 21 neuronal superclusters (13 excitatory, seven inhibitory, and one mixed neuronal supercluster). The supercluster labels capture major features but, inevitably for a complex tissue, some labels do not capture all features: for instance, “medium spiny neurons’’ and “eccentric medium spiny neurons” also contain cells from outside caudate and putamen (e.g., other long-range projecting inhibitory cells) and “amygdala excitatory neurons” also contain cells from paleocortex. Non-neuronal superclusters generally derived from dissections across the brain: e.g., astrocyte, ependymal, fibroblast, oligodendrocyte, and vascular cells were identified in many anatomical regions. Neuronal superclusters usually had a main anatomical region: e.g., deep-layer near-projecting and upper-layer intratelencephalic excitatory neurons were from neocortical dissections and the three hippocampal cell classes were from hippocampus (the chief exceptions were the more heterogeneous miscellaneous and splatter superclusters).

We identified protein-coding genes whose expression was highly specific for each brain cell type as assessed by top decile expression proportion (TDEP or “gene specificity”). Li et al. (in preparation) determined that the TDEP approach combined with S-LDSC had power and false positive rates that were jointly equivalent or superior to eight other methods (*Methods*). We also compared gene selection using relative versus absolute expression (i.e., TDEP versus TPM, transcripts per million), and found that both yielded nearly identical high-dimensional visualizations (*Figure S3*) but TPM was strongly influenced by broadly expressed “housekeeping genes” and this altered (and in some instances biased) gene ontology (GO) gene set analysis results (*Methods*).

**Figure 3.**
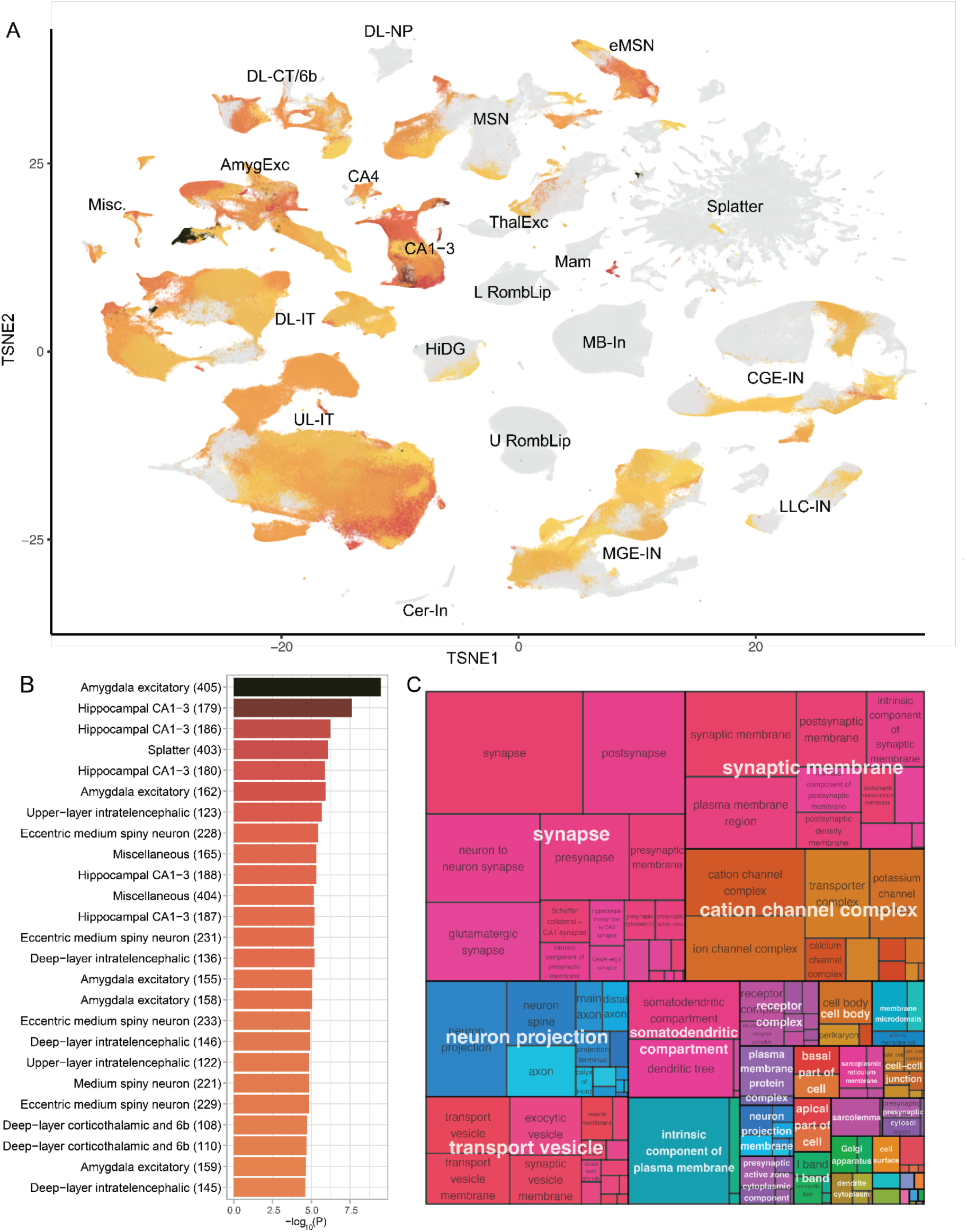
Cluster-level SNP-heritability enrichment for schizophrenia (scz2022). (A) tSNE plot from Siletti et al. colored by the significance of cluster-level SNP-heritability enrichment for scz2022. Gray indicates non-significance (FDR > 0.05). Abbreviations correspond to supercluster names in Figure 2A. AmygExc: Amygdala excitatory. DL-CT/6b: Deep-layer corticothalamic and 6b. DL-IT: Deep-layer intratelencephalic. DL-NP: Deep-layer near-projecting. CA1-3: Hippocampal CA1-3. CA4: Hippocampal CA4. HiDG: Hippocampal dentate gyrus. L RombLip: Lower rhombic lip. Mam: Mammillary body. Misc.: Miscellaneous. ThalExc: Thalamic excitatory. U RombLip: Upper rhombic lip. UL-IT: Upper-layer intratelencephalic. Cer-In: Cerebellar inhibitory. CGE-IN: CGE interneuron. eMSN: Eccentric medium spiny neuron. LLC-IN: LAMP5-LHX6 and Chandelier. MSC: Medium spiny neuron. MGE-IN: MGE interneuron. MB-In: Midbrain-derived inhibitory. (B) Top 25 significant clusters with the greatest scz2022 SNP-heritability enrichment, color as in Figure 2A. (C) Treemap plot for key GO-CC pathways in the top 25 scz2022 clusters. We performed gene set enrichment analysis for the TDEP genes in each of the top 25 significant clusters. Significantly enriched pathways in all the explored clusters were integrated to highlight higher level functions. The treemap for GO-BP and GO-MF for these clusters are shown in Figure S7.

We posit that TDEP genes for cell types are enriched for biological processes related to cellular identity and function. First, TDEP genes for different cell types generally had low overlap (*Figure S6*, median Jaccard index 0.049, IQR 0.022 - 0.099) but certain pairs had greater overlap (necessitating conditional analyses, *Figure 2A*). Second, as expected, “housekeeping” genes (highly and consistently expressed across tissues) ^34^ were markedly less likely to be TDEP genes. Third, we conducted GO gene set analyses ^35^ for lists of the ∼1,300 TDEP genes per supercluster (*Table S2*). Results for non-neuronal cells suggested a diverse range of significant GO terms consistent with the supercluster labels: astrocyte with biological adhesion, choroid plexus with cilium, microglia with immune response, oligodendrocyte with neuron ensheathment, oligodendrocyte precursor with gliogenesis, and vascular with vasculature development. GO terms for most neuronal cell types were dominated by synaptic biology, consistent with findings that forebrain neuronal cell identity is to a large extent driven by specific expression of synaptic genes ^36^ (exceptions were lower/upper rhombic lip and cerebellar inhibitory neurons from non-cortical regions). Fourth, the *Methods* section describes additional features of TDEP genes: (a) visualization of supercluster TDEP genes yielded groups of non-neuronal cells, neocortical excitatory neurons plus medium spiny neurons, and inhibitory interneurons plus non-cortical excitatory interneurons (*Figure S4A*); (b) TDEP genes tend to co-occur in genomic regions (*Figure S5*); (c) visualizations for gene expression specificity and genomic co-occurrence were similar suggesting that TDEP genes tend to be located near each other; and (d) all neuronal TDEP genes accounted for 61-65% of the SNP-heritability for the largest brain trait GWAS (scz2020, bip2021, mdd2019*, neuroticism, education, and IQ).

### Identifying human brain cell types implicated by GWAS

For each of the 36 primary GWAS, we estimated SNP-heritability enrichment for TDEP genes in each of the 31 superclusters (1,116 estimates). We used FDR correction for multiple comparisons per GWAS (*Figure 2A*, *Table S3*). The P-values were not uniformly distributed (modes near 0 and 1) and 9.6% of all comparisons had FDR < 0.05. Of the non-neuronal superclusters, only microglia reached significance for any trait (lymphocyte count, neutrophil count, and multiple sclerosis). As in our prior reports ^17,18^, non-neuronal superclusters were not significant for psychiatric disorders. In contrast, eight neuronal superclusters accounted for 60% of all significant enrichments. The numbers of superclusters with significant trait SNP-heritability enrichments were highly variable: (a) none of the structural MRI measures; (b) most neurological diseases had none (except for multiple sclerosis and epilepsy); (c) of the control traits, neutrophil and lymphocyte counts enriched for microglia and BMI enriched for two deep layer pyramidal cell superclusters and amygdalar excitatory neurons; (d) broad neocortical and non-cortical signals are also observed for other brain traits including educational attainment, IQ, neuroticism, and alcohol drinks per week with all neuronal neocortical superclusters significant for scz2022 and bip2021 (except for deep-layer near projecting). Non-cortical forebrain clusters from “amygdala excitatory” to “eccentric medium spiny neuron” showed strong signals for both scz2022 and bip202 (*Figure 2A*); and (e) the significant enrichments were dominated by six complex psychiatric disorder/brain traits. For the largest and most powerful GWAS traits (scz2022, bip2021, mdd2019, neuroticism, education, and IQ), the same eight neuronal superclusters had significant enrichment for all six traits. This agrees with the observation that schizophrenia, bipolar disorder, and MDD account for substantial morbidity and mortality and have considerable clinical and pharmacotherapeutic overlap (especially for severe and enduring forms of illness). Moreover, IQ, educational attainment, and the “Big 5” personality trait of neuroticism are important patient stratifiers and/or modifiers of clinical course ^37–39^. The neuronal superclusters were five excitatory (amygdala excitatory, deep-layer intratelencephalic, hippocampal CA1-3, hippocampal CA4, and upper-layer intratelencephalic) and three inhibitory (CGE interneuron, eccentric medium spiny neuron, and LAMP5-LHX6 and chandelier).

#### Excluding alternative explanations

We evaluated a set of potential explanations for the observed overlap of eight superclusters with six GWAS traits. First, the genome-wide genetic correlations between the six traits were occasionally high but far from complete: for the 15 unique genetic correlations, the median |r_g_| was 0.22, IQR 0.15-0.39. The largest r_g_ values were 0.73 for educational attainment-IQ, 0.69 for mdd2019-neuroticism, and 0.68 for bip2021-scz2022, and all other values were < 0.5. Second, the significant GWAS loci for these six traits only infrequently intersected with GWAS loci of more than one trait (median Jaccard index 0.069, IQR 0.034-0.094). Third, TDEP genes did not have fully explanatory overlap: of the 4,812 TDEP genes for the eight superclusters, 92.2% were TDEP for one (43.8%), two (24.7%), three (14.3%), or four (9.4%) superclusters. Fourth, we identified intersections of supercluster TDEP genes with GWAS loci, and found that TDEP genes (±50kb around each gene) only infrequently intersected more than one GWAS locus (7.6%). Finally and most directly, we conducted conditional analyses to evaluate the independence of the supercluster signals (*Figure 2B* and *Tables S4-S5*). Briefly, we found relatively consistent patterns of independence: amygdala excitatory, deep-layer intratelencephalic, hippocampal CA1-3, CGE interneuron, and eccentric medium spiny neuron generally survived conditional analyses suggesting the independence of most of the SNP-heritability results. Thus, we could identify no alternative statistical explanation or dataset redundancy to explain away the observed overlaps in *Figure 2A*.

#### Clinical subtyping

MDD has the advantage of large GWAS on its clinical subtypes. For instance, SNP-heritability estimated from GWAS where cases are people with severe MDD receiving electroconvulsive therapy is greater than when estimated from GWAS where cases are identified by self-report, community, or outpatient sampling ^41^. We compared the cell type enrichment between MDD subtypes viewed as clinically important (e.g., recurrent) or with empirical demonstration of greater heritability (e.g., highly severe MDD) with their counterparts ^41–44^. Categories with any significant superclusters are presented in *Figure 2C*, including recurrent MDD, MDD with functional impairment, MDD with suicidal thoughts, and postpartum MDD. In general, more signals were found in the more severe subtypes. Although the MDD subtypes largely overlap with mdd2019, there were indications of specificity; e.g., hippocampal superclusters are more related to the severe/impaired MDD subtypes and neocortical superclusters are more related to MDD with suicidal thoughts. In Table S1 we provide the GWAS sample size and the number of genome-wide significant loci (both benchmarks of GWAS power) for all traits, and note that the MDD subtype GWAS are relatively underpowered.

<u>Evolutionary constraint</u> has been of considerable interest given that SNP-heritability is notably enriched for this SNP annotation ^1^. *Figure 2D* depicts the distributions of a gene-based measure of constraint for TDEP genes for superclusters (the fraction of all CDS bases under strong constraint in 240 eutherian mammals). TDEP genes for inhibitory superclusters are more constrained than those for excitatory superclusters (in line with previous reports) ^45,46^. Schizophrenia has notable enrichment in evolutionary constrained genomic loci ^1,11^ (*Figure 2D*).

### Exome sequencing and neurodevelopmental disorders

*Table S6* presents analyses of supercluster TDEP genes where we evaluated gene annotations derived from LD-independent methods (e.g., whole exome sequencing). As a check, we found that TDEP genes for all superclusters were significantly less likely to be “housekeeping” genes, as expected given the definition of TDEP. There were no significant associations of any supercluster TDEP genes with genes implicated via whole exome sequencing for autism or schizophrenia ^13,47^, but there were many associations for developmental delay and neurodevelopmental disorder (NDD) ^47^. As the pattern of results was similar, we focused on NDD. NDD was significantly associated with TDEP genes for 15 of the 31 superclusters: (a) there were negative associations with Ependymal, Microglia, and Vascular superclusters (i.e., genes implicated in NDD were less likely to be TDEP genes); (b) there were eight significant associations with excitatory neuron superclusters (Amygdala excitatory, Deep-layer corticothalamic and 6b, Deep-layer intratelencephalic, Hippocampal CA1-3, Hippocampal CA4, Hippocampal dentate gyrus, Miscellaneous, and Upper-layer intratelencephalic); and (c) there were three significant associations with inhibitory neurons (Eccentric medium spiny neuron, LAMP5-LHX6 and Chandelier, and Midbrain-derived inhibitory). Notably, there was strong overlap of the NDD exome findings with the SNP-heritability enrichment for schizophrenia: of neuronal associations for NDD, 11 of 12 were also significant for schizophrenia and, of the associations with schizophrenia but not NDD, three of four were inhibitory neuronal superclusters. Clinically, a subset of people with schizophrenia have earlier NDD, and these results suggested that the two disorders may have important commonalities at a cell class level.

### Brain cell types implicated for schizophrenia

Siletti et al. ^27^ also identified 461 clusters of cells; superclusters contained a median of 12 clusters (IQR 8-17, ranging from 1 for Bergman glia to 92 for Splatter neurons). We conducted TDEP/S-LDSC analyses at the cluster level for schizophrenia (*Tables S7-S8*). The P-value distribution again had modes near 0 and 1. Of the 461 clusters, 199 (43.2%) had significant (FDR < 0.05) SNP-heritability enrichment for schizophrenia. There was a strong relationship of the SNP-heritability enrichment for schizophrenia in superclusters and their component clusters with most of the significant clusters in a significant supercluster (95.0%, 189/199). Of the 10 clusters not in a significant supercluster, Splatter 403 (GABAergic cells expressing *NOS1* from amygdalar and paleocortical dissections) was exceptional (FDR 9.6e-5) and the rest had FDR values between 0.009–0.05 (eight neuronal clusters and one non-neuronal cluster with FDR = 0.04). We again find little common-variant genetic support for non-neuronal cells in schizophrenia.

In *Figure 3A*, we visualized the supercluster and cluster findings for schizophrenia in the tSNE projection from Siletti et al. (their Figure 1B). The uneven distribution of schizophrenia associations across superclusters is readily apparent. Most of the 25 strongest cluster associations (FDR < 5e-4) were from a few superclusters (Hippocampal CA1-3, Amygdala excitatory, and Eccentric medium spiny neuron; Figure 3B). These clusters had a median of 1,274 TDEP genes (IQR 1,258–1,286) but with modest overlap between clusters (66.2% of unique genes were TDEP for ≤ 5 clusters). Gene set analysis of the TDEP genes in these clusters highlights synaptic function, cation channels, and neuron projection (Figure 3C, *S7*).

### Analysis of anatomic regions shows distributed risk for schizophrenia risk across the brain

Connecting genetic risk to specific brain regions is important for imaging (structural or functional MRI or PET) and for identifying empirical targets for Interventional Psychiatry therapeutics (e.g., transcranial magnetic stimulation). We evaluated the distributions of cell clusters across 104 dissections (2 dissections removed, *Methods*) from 10 broad brain regions (*Figure 4A*) ^27^. Neuronal clusters tended to be dissection-specific whereas non-neuronal clusters were widely distributed. As we observed a lack of signal in non-neuronal cell classes for psychiatric disorders (*Table S8*), and to avoid bias from different neuron/glia composition ratios, we focused on neuronal clusters. To evaluate SNP-heritability enrichment for anatomic regions, we computed the neuronal cluster proportions per anatomical dissection as weights for the cluster-level enrichments; the sum of the weighted cluster-level enrichment was the enrichment per anatomical dissection (*Methods*). First, we observed general effects in the cerebral cortex for scz2022, bip2021, educational attainment, IQ, and neuroticism (*Figure 4B*). Hippocampal, amygdala, and striatal (Pu and CaB) regions were also significantly enriched for the SNP-heritability of these phenotypes but with greater variability. Regional differences in hippocampal enrichment is consistent with analyses in mouse brain ^19^. Basal forebrain, thalamic, hypothalamic, cerebellum, and pons were enriched to lesser extents. To illustrate the distribution of genetic risk for schizophrenia across the brain, we depict the results using a 3D brain model (*Figure 4C-E*). Hippocampus and amygdala showed the highest significance of scz2022 SNP-heritability enrichment, with the top signal in the tail of hippocampus and the cortical amygdala (CoA). A detailed view of the neuronal cell type composition of the hippocampus and amygdala (*Figure 4F-G*) reveals that excitatory neuronal signals were the primary contributor to the hippocampal results. For amygdala, although the highest enrichment was found in the excitatory neurons, the inhibitory neurons had greater proportions and more significant enrichments than in the hippocampus (i.e., eccentric medium spiny neuron clusters).

**Figure 4.**
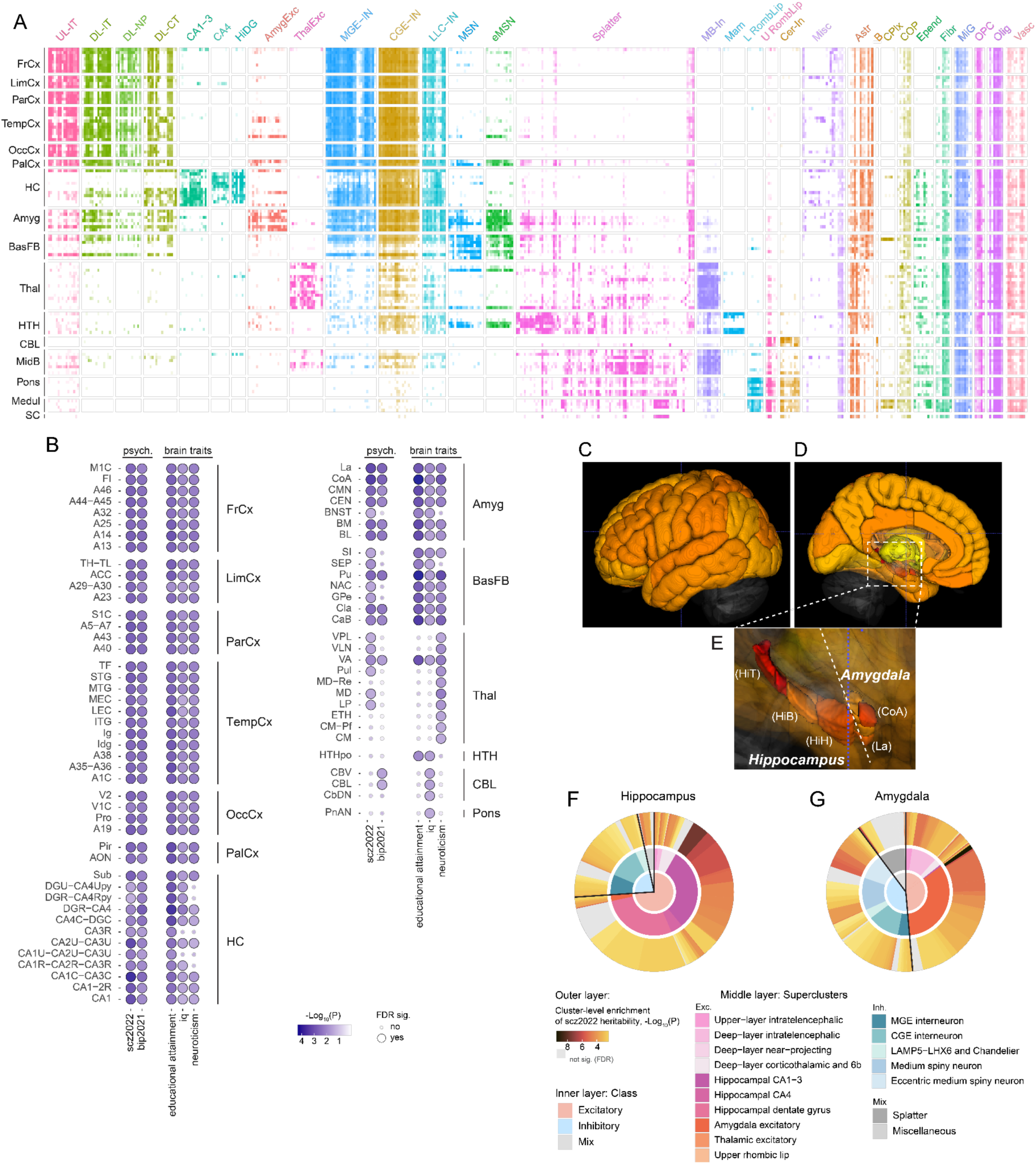
From clusters to anatomic dissections. (A) Heatmap of the scaled proportion of each cluster in each dissection. X-axis=clusters grouped and colored by superclusters. Abbreviations correspond to supercluster names in Figure 2A; neuronal abbreviations are the same as Figure 3A; non-neuronal abbreviations: Astr: Astrocyte. B: Bergmann glia. CPlx: Choroid plexus. COP: Committed oligodendrocyte precursor. Epend: Ependymal. Fibr: Fibroblast. MiG: Microglia. OPC: Oligodendrocyte precursor. Olig: Oligodendrocyte. Vasc: Vascular. Y-axis=brain anatomical dissections grouped to broader regions. Abbreviations: FrCx: frontal cortex, LimCx: limbic cortex, ParCx: parietal cortex, TempCx: temporal cortex, OccCx: occipital cortex, PalCx: paleocortex, HC: hippocampus, Amyg: amygdala, BasFB: basal forebrain, Thal: thalamus, HTH: hypothalamus, CBL: cerebellum, MidB: midbrain, Medul: medulla, SC: spinal cord. Each cell represents the scaled proportion of a cluster in a dissection. The proportions are the number of cells per cluster in a dissection divided by the total number of cells in the dissection; this number was then scaled to deciles for presentation clarity (Table S9). Nomenclature follows Siletti et al. 27. (B) Significance of S-LDSC SNP-heritability enrichment for anatomical dissections. The significance of enrichment at this was derived from cluster-level significance using the weighted-sum approach described in Methods. Dot color indicates level of significance in -log10(P), and darker blues indicate greater significance; dot size indicates the significance at FDR ≤ 0.05. Only phenotypes and brain regions with any significant signal are shown (Table S10). (C-E) Anatomic dissection results of scz2022 plotted on a 3D brain model (C-lateral view, D-sagittal view, E-enlargement of hippocampus and amygdala). Red indicates greater and yellow lesser significance at FDR ≤ 0.05, and gray and transparent indicates non-significance. Unsampled cerebral cortical regions are colored per the sampled regions (as mean of the enrichment Z-scores for sampled cerebral cortical regions). HiH: head of the hippocampus; HiB: body of the hippocampus; HiT: tail of the hippocampus; CoA: anterior cortical nucleus of the amygdala; La: lateral nucleus of the amygdala. (F-G) Greater detail for hippocampus and amygdala. The outer layer indicates clusters; the size is the proportion of the cluster, and the color indicates cluster-level significance of scz2022 SNP-heritability enrichment (color scale as in Figure 2B). The clusters are organized by the superclusters and sorted by the enrichment significance clockwise. The middle layer is colored by superclusters, and the inner layer is colored by classes. Splatter and Miscellaneous have both excitatory and inhibitory components and were categorized as “Mix”.

### Connectivity differences for hippocampus, amygdala, and cerebral cortex in schizophrenia cases

Although the prefrontal cortex, due to clear differences in those with schizophrenia, is the most studied brain region in schizophrenia, the cerebral cortex had consistently significant SNP-heritability enrichment. This, together with the prefrontal cortex having extensive connectivity with amygdala and hippocampus, suggested difference in the functional connectivity between the regions could contribute to schizophrenia mechanism, which we investigated using resting-state fMRI data from 46 cases with schizophrenia and 46 neurotypical controls (*Methods*) ^48^. We initially prioritized 76 brain regions that were enriched of the schizophrenia SNP-heritability (FDR ≤ 0.01) (*Table S11, Methods*).

We applied a deep neural network classifier to prioritize brain networks that distinguish cases from controls (*Figure 5A*). We randomly split the sample into five independent parts and performed five folds of parallel analyses, with four parts as the training set and one part as the test set (*Figure 5A*). We then performed recursive feature elimination such that the region with the lowest contribution was eliminated in the next iteration. Despite the limited sample size, four of five folds showed an upward trend for AUC (*Figure S8A, Table S12*), which is in line with the expected performance of the feature elimination process. At similar network sizes, these data-driven networks performed as well as or better than previously established, schizophrenia-relevant brain networks (i.e., the default mode and core networks, *Figure S8A*) ^28^. Hippocampal and amygdalar regions presented more frequently in the models with AUC>0.5 (*Figure 5B; Figure S8D*), and they were also enriched in regions with high numbers of connections (*Figure 5C*). To determine the most important connections across all the models in the five folds, we processed the feature importance of each pairwise connection (*Methods, Table S13*) and plotted the regions containing the top 0.5% (n=14) connections. Of the prioritized regions, three were from the hippocampus (left body, right body, and right head of hippocampus) and four were from the amygdala (left central nuclear group and the left lateral nucleus). Other prioritized regions, included the frontal lobe (posterior intermediate orbital gyrus, left middle frontal gyrus, and right rostral gyrus), right anterior cingulate gyrus, and parietal operculum (*Figure 5D*).

**Figure 5.**
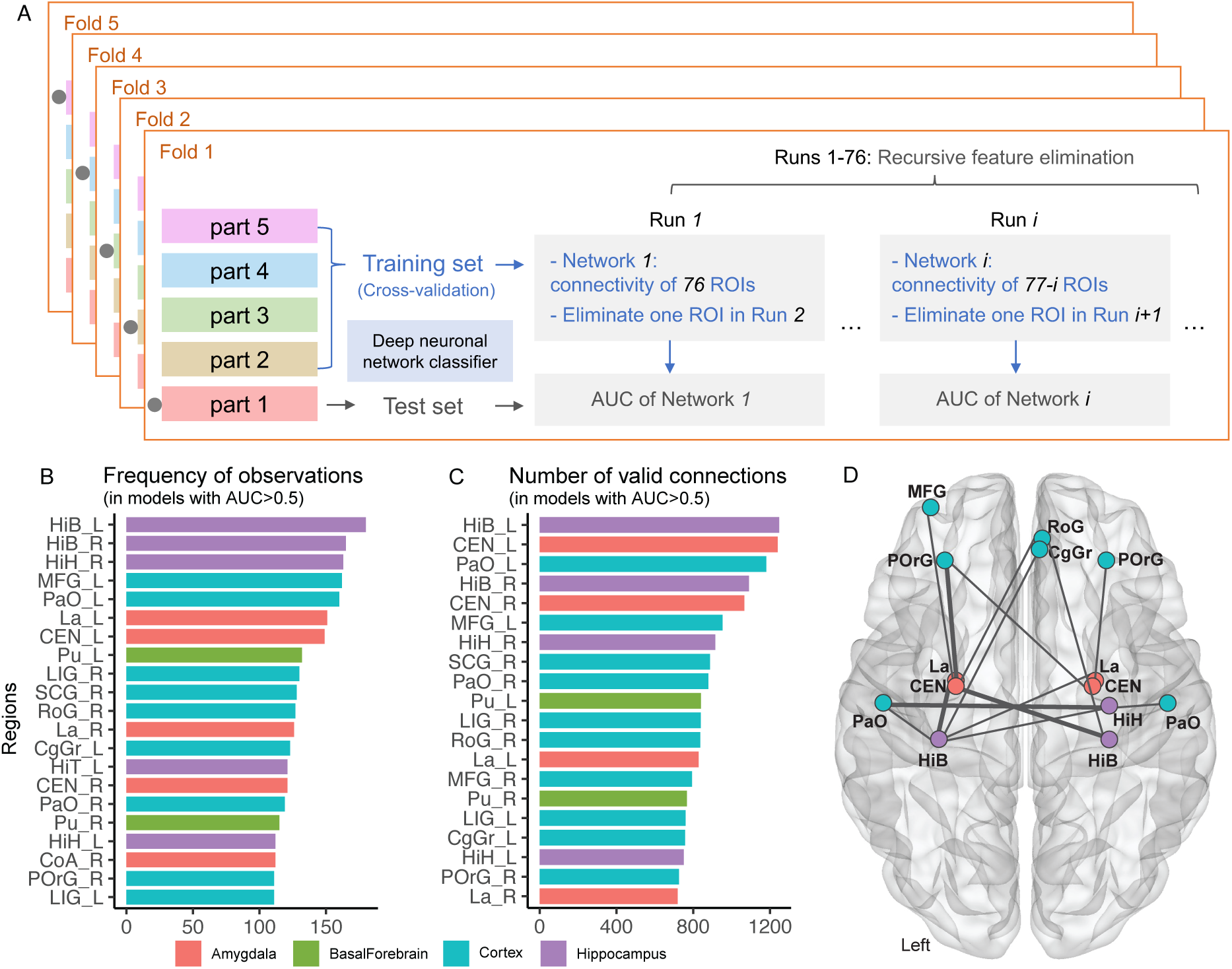
Functional connectivity networks derived from brain regions enriched for schizophrenia SNP-heritability distinguished cases and controls. (A) Workflow of the fMRI analysis, detailed in Method section. The gray dot to the left side of each portion indicates that the corresponding portion was used as the test set in the fold and that the rest were used as the training set. For each fold, the test set has always been kept separate from the training set, and across the folds, the test sets were independent from each other. (B) The top 20 regions that were most frequently observed in all the data-driven networks/models with AUC>0.5 across all folds. (C) The top 20 regions with the highest number of valid connections in the data-driven networks/models with AUC>0.5 across all folds. (D) Top 0.5% (n=14) connections across the data-driven networks/models with AUC>0.5 across all folds. Thicker segments indicate the top 4 connections/edges with the final edge strength>20; the rest connections/edges were marked with the thinner segments.

## Discussion

Psychiatric genomics now has empirical data strongly supporting polygenicity: multiple risk variants in “many genes” underlie the inherited tendency of these psychiatric disorders to run in families. The “genetic architectures” ^7^ of schizophrenia, bipolar disorder, MDD, and other major psychiatric disorders – causes of considerable human suffering – are dominated by large numbers of common genetic variants of small effect ^11,49,50^. The neurobiological implications of these secure and replicated genome findings are, however, unclear. In this paper, we rigorously evaluated the hypothesis that the accumulated findings implicate physically identifiable brain structures (i.e., cell types and anatomical regions). By necessity, our prior work was based on mouse brain atlases ^17,18^ and here we extend our work using a detailed and comprehensive human brain atlas ^27^. In a data-driven model, we show that the functional connectivity network inferred from genetically implicated brain regions had increased capacity to distinguish schizophrenia cases from controls compared to previously defined networks. This finding suggests a potentially important convergence between genomic findings and functional connectivity.

### Human brain cell data with regional resolution

Consistent with previous reports ^17,18^, neuronal cell types had substantially increased SNP-heritability for psychiatric disorders (schizophrenia, bipolar disorder, and major depression) and brain traits (educational attainment, iq, neuroticism, insomnia, alcohol consumption, and smoking initiation). The anatomical data allowed detection of trait-relevant brain regions. Regional signals were distributed across the cerebral cortex and subcortical cerebral nuclei. While confirming previous results based on mouse scRNA-seq data for hippocampal and neocortical excitatory neurons ^17,18^, based on human data we have identified novel relevant cell types, such as amygdala excitatory neurons, which were the most significantly enriched cell type in the entire brain as well as subcortical projecting GABAergic neurons for schizophrenia, which were undistinguishable in previous mouse datasets. Our study highlights neocortical interneurons derived from caudal ganglionic eminence which mainly contact other interneurons rather than interneurons expressing somatostatin or parvalbumin (although alterations of both of the latter have been reported in schizophrenia cases ^51,52^).

### Cross-disorder findings

We observed broad involvement of brain regions in several psychiatric disorders and brain traits; at the same time, each phenotype had multiple supercluster-level signals. These signals were largely statistically independent (*Figure 2B, Table S4-5*), suggesting different mechanistic contributions to the same cell types. Surprisingly, genes implicated by exome sequencing in neurodevelopmental disorders largely pointed at the same brain regions. Combined with our analyses of TDEP genes and GWAS loci, we believe that these results support cell types as contributing to phenotypically diverse traits. This suggests convergence, that these clinically distinctive phenotypes are rooted in different functional aspects of the same brain cell types.

With the available GWAS for MDD subtypes, we were able to infer important cell types for clinical subtypes. We observed that more superclusters were implicated for severer subtypes of MDD. It is possible that more severe subtypes convey higher genetic risk and therefore greater statistical power in the GWAS. It is also possible that the different subtypes had partially distinct etiologies, as suggested by imperfect genetic correlations among subtypes ^44,53^. More precise interpretations of the cell types can be made when larger subtype GWAS become available.

At the level of brain regions, the results pointed out the importance of subcortical structures, especially the hippocampus and amygdala, underlying the mechanisms of pathological (e.g., schizophrenia) and healthy (e.g., educational attainment) phenotypes. The results in amygdala are in agreement with other findings that implicates changes in its structure in psychiatric disorders ^54–56^. From a clinical perspective, amygdalar dysfunction agrees with decreased ability to ascribe correct valence and attention to sensory inputs ^57^.

### Implications for schizophrenia

Neocortical regions presented similar enrichments across the brain even though certain neocortical regions have been implicated in psychiatric disorders (e.g., dorsolateral prefrontal cortex and schizophrenia ^58^). This is likely explained by the similar cell type composition across the neocortical regions ^59^, and highlights the importance of functional connectivity in the underlying mechanisms of schizophrenia ^60^. The TDEP genes of the top scz2022 clusters highlighted synaptic functions and neuronal projection suggesting mechanistic connectivity between cells.

fMRI connectivity networks that we constructed from the brain regions enriched for schizophrenia heritability were both similar (DMN) and different (Core Network) between cases and controls to those previously identified as schizophrenia-relevant ^28^. Subcortical regions, although seldom studied together with neocortical regions in defined brain networks ^61,62^, were highlighted as critical contributors in our data-driven, genetically implicated brain networks. Resting-state connectivity of subcortical structures has received relatively little attention in schizophrenia, although they connect to the neocortical regions and demonstrate a similar level of complexity ^63,64^. Nevertheless, disturbed connection between the amygdala and the ventral prefrontal and the nearby orbitofrontal cortices has been reported in independent studies ^65,66^. Across our data-driven models distinguishing between patients and controls, the hippocampal and amygdalar structures possessed connectivities between each other and to cortical regions. Both hippocampus and amygdala are involved in emotional memory processing, and a directed influence from the amygdala on the hippocampus has been suggested during fear processing in response to emotionally salient information ^67,68^. Our method thus suggests reasonable brain regions and networks for schizophrenia etiology and calls for further investigations into these areas and their connectivities, which may hold new candidates for modulation using non-invasive therapeutics. Taken together, this study shows how genetic findings, combined with single-cell transcriptomics, can be used to prioritize not only cell types but brain regions and that these can be linked to disease relevant changes in functional connectivity. Our approach provides hope that the two modalities of genetics and brain imaging eventually are pointing towards the same targets.

These results need to be considered with limitations (see also the Supplement of reference ^17^). First, brain regions were not equally sampled (*Table S14*), despite the snRNA-seq dataset having the most comprehensive coverage of the adult human brain to date, and we cannot rule out enrichments of trait heritabilities in other brain regions. Second, the Human Brain Atlat is from a few adults and does not capture variability between neurotypical individuals or individuals with severe and enduring mental disorders or variability across the lifespan (especially during brain development). Third, TDEP is a relative measure that depends on which cell types are included. It is best applied in a comprehensive cell type atlas like the Human Brain Atlas and caution is warranted when comparing results from different cell type databases. Finally, we were unable to account for bilaterality given that snRNA-seq data were from the right hemisphere and the fMRI data were bilateral.

In conclusion, our findings extend prior work by showing the human brain localization of genomic regions implicated in three psychiatric disorders, three relevant brain traits, and in genes implicated in neurodevelopmental disorders. The findings point at largely overlapping cell types and brain regions (albeit different subsets of genes). These findings provide a framework for understanding the polygenicity of complex psychiatric disorders and brain traits as well as suggesting hypotheses for future research. Our findings underscore the value of single-cell transcriptomics in decoding the polygenicity of psychiatric disorders and offer a promising convergence of genomic, transcriptomic, and brain imaging modalities toward common biological targets.

## Methods

### Human Brain Atlas single-nucleus RNA-seq (snRNAseq)

We used the Human Brain Atlas snRNAseq data set from Siletti et al. ^27^. This atlas consists of 3.369 million nuclei successfully sequenced using snRNAseq. The nuclei were from adult postmortem donors, and the dissections focused on 106 anatomical locations within 10 brain regions. Following quality control, the nuclear gene expression patterns allowed the identification of a hierarchy of cell types that were organized into 31 superclusters and 461 clusters. In the current paper we use the same naming system for the cell types and the brain regions as in Siletti et al.

### Genome reference and gene models

The reference genome and gene models were with respect to a modified version ^27^ of the GENCODE primary assembly (GRCh38.p13, v35, 3/2020, hg38) ^69^. As hg19 is typically used by GWAS, we also obtained GRCh37/hg19 gene coordinates from GENCODE (v35). In these analyses, we focused on 18,090 genes with these characteristics: protein-coding, mapped to canonical autosomes (chr1 to chr22), not in the extended major histocompatibility (MHC) region (chr6:25-34 mb), and expressed in ≥ 1 of the 461 cell clusters. Explanations for these choices follow.

#### Protein-coding biotype

The modified GENCODE assembly used by Siletti et al. ^27^ contained N=51,263 genes with TPM > 1 in one or more cluster cell types. In GENCODE, these genes are grouped into 30 biotypes ranging from rare (“scRNA” and “vault_RNA”) to common (“protein_coding” (N=19,153) and lncRNA (16,021)). Siletti et al. used the 10X Genomics Chromium Next GEM Single Cell 3’ Reagent Kits (v3) whose beads contain a 30 nt poly-dT tail and thus will most consistently capture 3’ poly-adenylated RNA transcripts (in humans, these include mature protein-coding and lncRNA transcripts). For each biotype, we summed the number of occurrences of any gene with TPM > 1 over all superclusters and found that only protein-coding and lncRNA genes had appreciable transcript detection. For instance, 20 biotypes had < 100 detected transcripts and 28 had < 8,600 detected transcripts in any supercluster. We chose to drop lncRNA genes and only include protein-coding genes. First, although a small number of lncRNAs have been shown to have biological functions, the annotation of most lncRNAs is currently unknown. In these data, 81.4% of the lncRNAs had a generic annotation (e.g., “novel transcript”). Second, the lncRNA were not strongly expressed and/or were not well-captured by the 10X Genomics kit: the largest median expression of lncRNAs in the superclusters was only 0.20 TPM (compared to 11.5 TPM for protein-coding genes).

#### MHC

The extended MHC (eMHC) is the largest block (∼8 mb) of high linkage disequilibrium (LD) in the genome (excluding pericentromeric regions) ^70^. For instance, of the 23,731 significant SNP associations with schizophrenia, 4,527 (19.1%) are in the eMHC region ^11^. These generally correspond to highly correlated genetic variants. We removed GWAS SNPs and snRNAseq data in the eMHC as in our prior papers and as recommended by the S-LDSC authors ^17,18,71^. However, to evaluate the impact of this choice, we recalculated the TDEP estimates while including protein-coding eMHC expression data (N=259), and found that a small number of eMHC genes had a TDEP flag in superclusters (median 10 genes, IQR 8-13). As the median number of TDEP genes per supercluster was 1,287, the potential eMCH region contribution to a TDEP list is 0.8% (10/1287). The impact is likely not consequential.

#### Autosome

Sex chromosome genes were removed. chrY is rarely included in GWAS; in a recent build of the NHGRI/EBI GWAS Catalog ^72^, there were only 5 significant SNP associations to any trait whereas a similarly sized chromosome (chr22) had >3,000 GWAS hits. In addition, chrX data are inconsistently included in the summary statistics from GWAS papers ^73^, and are under-represented in the GWAS catalog: chrX has 1,149 hits whereas the similarly sized chr7 and chr8 have 9,438 and 9,745 associations. In a sense, the choice to exclude sex chromosomes was made for us as, for the GWAS traits we analyzed (*Table S1*), none had chrY and a minority had chrX results. To evaluate the impact of this choice, we recalculated the TDEP estimates while including protein-coding chrX expression data (N=779) and found that some chrX genes had a TDEP flag in superclusters (median 49 genes, IQR 41-60). As the median number of TDEP genes per supercluster was 1,287, 49 genes (3.8%) may have had a small impact. We are unable to address this issue given the data available, and this is unquestionably a topic for future research.

### GWAS summary statistics

We conducted multiple searches to identify potential GWAS (i.e., PubMed, Psychiatric Genomics Consortium downloads page, NHGRI/EBI GWAS catalog). We previously have shown the importance of genetic architecture on the informativeness of our approach (see ^11,42^. The number of loci (genomic regions harboring multiple correlated genome-wide significant SNPs, defined below) is particularly important. We required > 10 loci for inclusion (with a few intentional exceptions). *Table S1* summarizes the GWAS included in our analyses. These 36 primary GWAS are the largest studies per trait that we could obtain as of 4/2023 and whose use was compatible with our publication strategy (some prepublication GWAS required submission delays and others were not freely available, e.g. 23andMe).

● We included five psychiatric disorders (ADHD, bipolar disorder, major depressive disorder, problematic alcohol use, and schizophrenia). We did not include multiple important psychiatric disorders due to low numbers of loci (e.g., anorexia nervosa, autism).
● We included eight neurological diseases: Alzheimer’s disease, amyotrophic lateral sclerosis, epilepsy, hearing loss, migraine, multiple sclerosis, Parkinson’s disease, and stroke. For Parkinson’s disease, we used the results of Nalls et al. 2019 excluding 23andMe samples ^74^.
● We included nine structural brain MRI measurements: brainstem volume, caudate volume, neocortical surface area, and putamen volume. Because these MRI measures describe important brain features (and often the anatomic regions from Siletti et al.), we also included accumbens volume, amygdala volume, neocortical thickness, pallidum volume, and thalamus volume.
● We included nine trans-diagnostic brain traits of clinical salience (alcohol use, smoking traits, and insomnia) or which may be clinical stratifiers (educational attainment, IQ, neuroticism). Suicide phenotypes were included due to their importance in the current mental health crisis.
● Finally, we selected five control traits with large numbers of loci but whose genetic architectures are not rooted in the central nervous system: height, body mass index, hematocrit, lymphocyte count, and neutrophil count.

For MDD, we included 19 additional GWAS to assess within-disorder questions (no requirement for minimum number of associations; *Table S1*). As etiological heterogeneity is likely for depressive disorders, we evaluated whether heterogeneity was associated with different brain cellular enrichments. We focused on the clinical contexts in which a major depressive episode (MDE) can occur. The classical delineation of MDE is in the context of unipolar or bipolar disorder. An MDE can occur as major depressive disorder (MDD, unipolar MDE with no history of mania or hypomania), MDE with a history of mania (bipolar disorder type 1), and MDE with a history of hypomania (bipolar disorder type 2). These conditions have different genetic correlations with bipolar type 2 being more similar to MDD and bipolar type 1 being more similar to schizophrenia ^49^. MDD can occur in different ways clinically and across the lifespan. We evaluated MDD subtypes viewed as clinically important (degree of severity, typical vs atypical symptom pattern, with or without comorbid anxiety disorder) or with empirical demonstration of greater heritability: highly severe MDD (people receiving electroconvulsive therapy for MDE), early-onset MDD, recurrent MDD, and postpartum depression ^41–44^.

#### Processing and quality control (QC)

After we obtained GWAS results from the primary sources, we conducted range checks for logistic or multiple regression betas, standard errors, and P-values (removing SNPs with highly unlikely values). We then processed all sumstats using the `cleansumstats` pipeline (https://github.com/BioPsyk/cleansumstats):

● We determined genome build by comparing SNP positions to dbSNP (build 151) ^75^
● Using `UCSC::liftOver`, we ensured we had sumstats in hg38/GRCh38 and hg19/GRCh37 coordinates (GWAS tend to use hg19 and genome annotations tend to use hg38)
● We removed insertion/deletion polymorphisms, duplicate entries, and chromosomal locations not in [chr1-chr22] and noting that sex chromosome data are inconsistently included in GWAS summary statistics ^73^
● We required that each variant match dbSNP (build 151) by rsID and that the GWAS sumstats SNP alleles (effect/other allele) matched REF/ALT in dbSNP (flipping to + strand if required)
● We removed homozygous/monomorphic SNPs, SNPs with alleles not in [ACGT], and strand-ambiguous SNPs (A/T or C/G; these are also removed in LD score regression)
● Given our use of S-LDSC (below, and as typically done), we excluded the extended MHC region (chr6:25-34 mb) due to its exceptionally high LD

#### Genomic loci

We used the clumping algorithm in `plink` ^76^ to identify loci for the GWAS included in this report. The LD reference was the European subset of the 1000 Genomes Project (phase 3) ^77^ with parameters: p1=5e-8, p2=5e-6, r^2^=0.1, and window size of 3,000 kb. Overlapping loci and loci within 50 kb of each other were merged.

#### Description of the primary GWAS

We then conducted basic checks including the number of SNPs after QC, the number of genome-wide significant SNPs (P < 5e-8, after QC), inflation statistics (𝜆 and LDSC intercept), and SNP-heritability (*Table S1*). For the primary GWAS traits, the numbers of significant loci were positively correlated with sample size (Spearman 𝜌 = 0.62, P = 4.9e-5) and the number of cases (binary traits, Spearman 𝜌 = 0.73, P = 0.0012). *Figure S1* illustrates some key features of the GWAS included in the primary analyses.

Figure S2 provides more data about the primary GWAS traits. The SNP-heritability estimates on the diagonal are consistent with the primary reports (any differences are due to our use of sample subsets like European subjects or after removing 23andMe results). The off-diagonal elements show the interrelationships of the primary GWAS traits via a heatmap of genetic correlations (rg from LDSC). The pattern of genetic correlations are consistent with prior reports ^3^. In general, we note: (a) positive intercorrelations for psychiatric disorders and brain traits, (b) transdiagnostic negative correlations of educational attainment and IQ with multiple conditions, (c) relatively weak correlations for neurological diseases, and (d) isolated correlations for structural MRI measures.

### Relative versus absolute gene expression

We use top decile of expression proportion (TDEP) to identify genes whose expression typifies each supercluster (∼1,300 genes per supercluster). Li et al. (in preparation) determined that S-LDSC with TDEP had power and false positive rates that, jointly, were equivalent or superior to 8 other methods. See the *Statistical analysis* section below for definition of TDEP, TPM, and the Li et al. results).

Here, we compare relative vs absolute measures of gene expression. TDEP is a relative measure, the expression of a gene in one cell type divided by the total expression across all cell types. In contrast, TPM (snRNAseq count data in a cell type normalized to molecule transcripts per million) is more of an absolute method that reflects the number of RNA molecules in specific cells. We thus contrasted TDEP and TPM. \

First, as a basic data visualization, *Figure S3* depicts the relation between TDEP and TPM for each of the 31 superclusters. For most superclusters, gene expression was greater in TDEP genes. This was particularly notable for non-neuronal superclusters where the median expression was far higher for TDEP genes (e.g., oligodendrocyte median 102.1 vs 18.9 TPM in TDEP genes vs all other genes). The excitatory neuronal superclusters had similar appearances in *Figure S3* except for upper rhombic lip and lower rhombic lip being somewhat different. Inhibitory neuronal superclusters appeared relatively similar.

Second, we created two data matrices; rows were 18,090 autosomal, protein-coding genes, columns were 31 supercluster classes, and the elements were either log_2_(TPM+1) or TDEP (1=yes, 0=no). The TPM matrix is obviously far more nuanced and detailed than the TDEP version). The results of UMAP/HDBSCAN are depicted in *Figure S4*. In both instances, the high-dimensional data could be visualized as 3 distinct clusters. Cluster positions are arbitrary but the solutions are otherwise qualitatively similar, clusters containing: (a) all non-neuronal cells; (b) all neocortical excitatory neurons plus medium spiny neurons; and (c) all inhibitory neurons and non-cortical excitatory neurons.

This is notable because TDEP faithfully recapitulates the multivariate structure of the supercluster data based on the more information-rich and full gene expression matrix based on TPM. The TDEP 0/1 flags efficiently capture the high-dimensional density structure of the TPM expression matrix.

Third, we contrasted gene set analyses using GO ^35^. The GO gene set analyses were based on TDEP genes and separately for top decile TPM. As both variables are defined by the deciles and coded TRUE/FALSE, similar numbers of genes are being compared. The background was 18,090 autosomal, protein-coding genes.

These analyses yielded 322,462 comparisons (31 superclusters x 10,422 GO sets). The correlation in hypergeometric P-values for TDEP with top decile TPM was modest (Spearman 𝜌 = 0.414). For significance at FDR < 0.05, TDEP was more conservative than top decile TPM in implicating GO gene sets (3.27% vs 8.40%). Of all pathways, the two methods agreed for 92.2% (both non-significant for 291,094 or 90.30%, and both significant for 6,228 or 1.93%). There were fewer disagreements for TDEP==TRUE and top decile TPM==FALSE (4,329 or 1.34%) than the reverse (TDEP==FALSE and top decile TPM==TRUE, 20,811 or 6.45%). Checks of disagreements with top decile TPM FDR < 0.05 and TDEP FDR > 0.5 (larger FDR applied to avoid edge cases) revealed some confusing results: e.g., synaptic genes sets with non-neuronal supercluster classes including astrocyte, Berman glia, oligodendrocyte, fibroblast, and vascular. These disagreements tended to be the same (i.e., about half of these pathways were implicated in ≥5 superclusters). We believe that the differences were strongly influenced by broadly (and often highly) expressed “housekeeping genes” that are prevalent in top decile TPM but not in TDEP (by definition). The top decile TPM gene set findings are in contrast to those for TDEP (presented in *Table S2*) that captured the expected (if not canonical) biological processes, cellular compartments, and molecular function of the 31 superclusters.

Taken together, these results support our use of TDEP as a means to identify genes that are enriched for biological processes related to the cellular identity and specific function of superclusters.

#### TDEP in human and mouse brain studies

Our prior papers were based on scRNA-seq mouse neural surveys ^17,18^ with the key limitation of a necessary reliance on protein-coding genes with a high confidence, 1:1 mouse-human ortholog. Of the 18,090 genes we evaluated (autosomal, protein-coding, not in eMHC, TPM > 1 in ≥ 1 cluster), 14,398 (79.7%) had a high confidence, 1:1 mouse-human ortholog. As a sanity check, we compared TDEP genes for 23 mouse brain cell types used in Skene et al. ^17^ to the 31 Human Brain Atlas supercluster using hypergeometric gene set analysis. There was considerable consistency across these datasets despite different technologies and organisms. For example, there was the greatest overlap of: mouse “pyramidal CA1” with human hippocampal CA1-3 (fdr = 1e-140); mouse “pyramidal somatosensory” with human upper-layer intratelencephalic (fdr = 1e-134); mouse “oligodendrocyte” with human oligodendrocyte (fdr = 1e-183); and mouse “endothelial mural” with human vascular (fdr < 2e-208). As expected, the mouse signal for some cell types resolved into more precise human superclusters: mouse “interneurons” was associated with four human inhibitory neurons and the three mouse hypothalamic cell types contained human ependymal as well as excitatory and inhibitory neuronal TDEP genes.

#### Summary

Thus, we believe that TDEP is a defensible choice. Its relative nature can be a limitation in extreme instances but it is a principled and intentional choice that we evaluated extensively in this section. Further support can be found in method comparison studies: see discussion of Li et al. in the section titled “Choice of TDEP/S-LDSC”. The similarities in *Figure S4* are reassuring and TDEP’s more conservative and the face-valid gene set results strengthen its appeal.

### Properties of Human Brain Atlas superclusters

We use TDEP to identify genes whose expression typifies each supercluster (∼1,300 genes per supercluster). Multiple choices that we made in using TDEP are explained above, and the *Statistical Analysis* section below provides definitions and further justification. We posit that TDEP genes for a cell type are enriched for biological processes related to cellular identity and function, and we evaluated this assumption in multiple ways.

#### Traditional classification and gene set analysis

*Table S2* characterizes the 31 supercluster cell classes: 10 non-neuronal and 21 neuronal cell classes (13 excitatory, 7 inhibitory, and 1 mixed neuronal class). Non-neuronal cell classes generally derived from dissections across the brain: e.g., astrocyte, ependymal, fibroblast, oligodendrocyte, and vascular cells were identified in many anatomical regions (with the exceptions of Bergmann glia and choroid plexus). Neuronal cell classes usually had a predominant anatomical region: e.g., deep-layer near-projecting and upper-layer intratelencephalic excitatory neurons from neocortical dissections and the 3 hippocampal cell classes were from hippocampus (the main exceptions were the miscellaneous and splatter).

*Table S2* contains Gene Ontology (GO) gene set analysis for TDEP genes ^35^. Results for non-neuronal cells suggested a markedly diverse range of significant GO terms that were consistent with the supercluster labels: astrocyte/biological adhesion, choroid plexus/cilium, microglia/immune response, oligodendrocyte/ensheathment of neurons, oligodendrocyte precursor/gliogenesis, and vascular/vasculature development. In contrast, for most neuronal cell classes, GO terms focused directly on synaptic biology.

A small set of genes (215, 1.19%) had TDEP in 10-14 supercluster classes. These genes contained multiple cadherins, calcium channel subunits, muscarinic receptors, GABA receptors, glutamate ionotropic and metabotropic receptors, potassium channel subunits, sodium channel subunits, synaptotagmins, and transmembrane proteins. Despite a small number of genes that usually limits gene set analysis, these 215 genes were enriched for 38 SynGO ^78^ synaptic cellular compartment and biological process annotations (e.g., presynapse P_hyper_ = 3.9e-11 and postsynapse P_hyper_ = 4.1e-11).

We also addressed the inverse question, the 21.0% of genes that were not in a TDEP gene list for any supercluster. These genes were highly enriched for: (a) genes expressed at high and consistent levels across tissues (P_hyper_ < 2.2e-308, a definition of “housekeeping” genes) ^34^; (b) evolutionarily constrained genes (P_hyper_ = 1.6e-108) ^1^; (c) a range of GO biological process annotations pertaining to RNA processing, gene regulation, and cellular energetics (P_hyper_ < 1e-40); and (d) notably, no synaptic processes (P_hyper_ = 1) ^78^. As expected, genes whose supercluster expression are non-specific were dominated by fundamental processes common to most cells and which tend to be highly constrained in placental mammals.

#### Gene expression

The Human Brain Atlas data ^27^ consist of snRNAseq on 3.369 million nuclei from adult postmortem donors and 106 anatomical locations within 10 brain regions that were then organized into 31 supercluster classes. We made a data frame with columns for the Ensembl gene identifier and each of the 31 superclusters along with 18,090 rows (for each autosomal, protein-coding gene expressed in ≥ 1 cluster). The elements are the expression of a gene in each cell class (as TPM, molecule transcripts per million). *Figure S3* shows the relation between TPM and EP by supercluster class. At this level of analysis, there is considerable diversity in terms of the gene repertoire and expression level. Many of these genes will be responsible for core physiological processes and are robustly expressed in most cells (e.g., “housekeeping” genes).

#### Genomic location and supercluster TDEP genes

Genes are not randomly positioned in the human genome but rather show a marked tendency to occur in clumps. For instance, we can divide the genome into a regular set of 100 kb bins. After removing bins that were entirely composed of “N” (unknown) bases and after excluding chrX, chrY, and the extended MHC region (as noted above), there were 27,597 x 100 kb bins. We assigned the transcription start site (TSS) of 18,090 protein coding genes to these 100 kb bins, and tabulated the observed number of bins with 0, 1, 2, … TSS. We created an expectation using random sampling with replacement. In the human genome, we observed that 97.1% of all 100 kb bins have no TSS (i.e., 2.9% contain from 1-14 TSS). The observation is markedly different from the random expectation: the fraction of bins without a TSS ranged from 78.3-80.0% (1,000 trials). As the observed fraction of bins with no TSS (97.1%) was never approached, this implies that an empirical probability of this observation is far less than 0.001.

If protein-coding genes clump or cluster together in the genome, then TDEP genes are likely to cluster as well, given that they are a subset of all protein-coding genes. We thus assessed whether TDEP genes co-occurred in excess of the fundamental clumping of protein-coding genes. For each of the 10,027 x 100 kb bins containing a TSS for a protein-coding gene, we tabulated the total number of protein-coding TSS (nTss) and the number of these that were TDEP genes (separately for each supercluster). We fit 31 linear regression models (nTdep**_i_** ∼ nTss) and saved the studentized residuals (i.e., transformed to mean 0 and standard deviation 1). The studentized residuals had minimum values > -3 and the 75th percentiles were around -0.2. However, for ∼14% of the bins, there were far more TDEP TSS than expected given the number of TSS (defined as studentized residuals > 3). To visualize these relationships, we computed the Spearman correlations for the studentized residuals of 31 supercluster cell classes and depicted the correlation matrix as a heatmap following hierarchical clustering (*Figure S5*). Note that: (a) non-neuronal supercluster classes tend to correlate, specifically ependymal-choroid plexus, Bergman glia-astrocyte, oligodendrocytes, and vascular-fibroblast; and (b) neuronal cells classes clump in 2 groupings.

The groupings in *Figure S5* are strongly reminiscent of those in *Figure S4*. We believe that this is notable given that the input data come from different sources (the latter from genomic location and the former from gene expression). This observation implies a role for co-expression of genes in genomic regions and supercluster identity.

### Statistical analysis for SNP-heritability enrichment

#### Gene expression specificity/expression proportion

We calculated gene expression specificity per cell type as expression proportions (EP). The following steps were done for cell types at the supercluster level (N = 31) and then at the cluster level (N = 461). For each cell type, we normalized the Siletti et al. ^27^ snRNAseq count data to molecule transcripts per million (TPM, equation I). We then computed EP per cell type as the normalized expression divided by the sum of normalized expression across cell types for each gene (equation II).

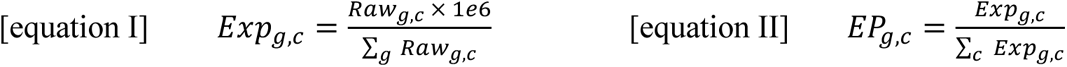

As in our prior papers, we selected genes in the top decile of expression proportion (TDEP) per cell type with normalized expression >1 TPM.

#### SNP-heritability enrichment in cell types

Stratified LD score regression (S-LDSC) is widely used to evaluate whether a specific genome annotation is enriched for GWAS findings that contribute a greater proportion of SNP-heritability (also known as SNP-based heritability) to the common variant genetic architecture. It incorporates an empirical approach to LD correction via LD scores (the sums of local *r*^2^ LD values) and includes multiple other genome features to increase model stability ^71,79^. We used S-LDSC ^79^ to evaluate whether the set of ∼1,300 genes with top decile EP for each supercluster-level cell type (N = 31) or cluster-level cell type (N = 461) had significant SNP-heritability enrichment. Enrichment is calculated for the SNP-set relative to a null hypothesis that all SNP contribute equally to the SNP-heritability. Gene boundaries were expanded by ±100 kb. S-LDSC was run for each combination of GWAS summary statistics and cell type (at supercluster- and cluster-levels). We provide additional justification for these methodological choices below. As recommended, enrichment P-values were computed from the “Coefficient_z-score” ^79^. For each GWAS trait, we adjusted for multiple comparisons using false discovery rate (FDR) using `R::rstatix::adjust_pvalue(method="fdr”)`.

#### Choice of TDEP/S-LDSC

Multiple groups have proposed algorithms by which to connect GWAS results to specific cell types. In addition to top-decile EP (TDEP)/S-LDSC, published methods include (in alphabetical order): `CELLEX`, `DIALOGUE`, `EPIC`, `EWCE`, `MAGMA`, `RolyPoly, sc-linker`, and `scDRS` ^19–26^. These methods evaluate the association of GWAS signals with gene expression specificity in a given cell type as measured by single-cell or single-nucleus RNAseq.

Co-authors Ang Li, Jian Zeng, and Naomi Wray (University of Queensland and Oxford University) have conducted a comparison of a representative set of these methods (manuscript in preparation). Broadly, the methods are based on SNP-level regression (e.g., LDSC), gene-level regression (e.g., MAGMA-set), and cell scoring methods (e.g., scDRS). Methods that use SNP-level or gene-level regression methods differ in their specificity metrics to determine gene sets per cell type from the RNAseq date before integration with GWAS summary statistics. In contrast, scDRS takes a set of associated genes for a trait from the GWAS summary statistics into analyses of the single-cell RNA-seq data. Li et al. compared the performance of 9 representative methods: the approach used here (TDEP/S-LDSC) ^17,18^, MAGMA-set + EP, scDRS (using MAGMA-gene to select the top 1,000 genes), sc-linker, and 5 CELLEX statistics (DET, GES, EP_w_, NSI, ES_µ_). They evaluated these methods with respect to empirical data using the default settings from each method: 18 GWAS trait/cell type pairs for which no association was expected (e.g., proerythroblast cell type and asthma) and 18 GWAS trait/cell type pairs for which there was independent evidence for association (e.g., proerythroblast cell type and red blood cell count). Application of these methods to empirical data sets allowed estimation of false positive control and power in real-world scenarios.

In brief, Li et al. found: (a) the method we use in this report (TDEP/S-LDSC) had power and false positive rates as good as or better than other methods; (b) MAGMA-set had somewhat higher power but at the cost of high false positive rates (indeed, we observed that MAGMA-set can yield markedly discrepant evidence for GWAS-cell type linkages – e.g., P-values 5-10 logs smaller than TDEP/S-LDSC); and (c) use of scDRS was constrained by computational burden – use of all 3.3 million cells for one GWAS trait took ∼40 compute hours and 600 gb memory on a high-performance Linux cluster so that applying scDRS to the 36 primary GWAS was infeasible without down-sampling to a subset of nuclei. In addition, other authors have noted that TDEP performs well with respect to other expression metrics (Appendix 2, Figure 3 in reference ^26^).

#### S-LDSC gene boundary expansion (±100 kb)

Expanding gene boundaries by ±100 kb is often done and is generally consistent with the locations of promoters, enhancers, and eQTLs that impact gene expression ^26,71^. We compared gene boundaries of ±100 kb and ±50 kb. For supercluster-level cell types and across the primary GWAS traits (*Table S1*), the correlation between log_10_(enrichment-P) for ±100 kb vs ±50 kb was 0.988. We also calculated Cohen’s kappa for the significance of the results (FDR correction) between the two choices of windows [ using `R::irr::kappa2()` ]. Even considering the conservative impact of significance thresholding, Cohen’s kappa between the two windows was as high as 0.93. Given the small differences for these two gene expansion windows and to remain consistent with our prior papers ^17,18^, we used gene boundaries ±100 kb.

#### Conditional analysis of gene specificity overlap

An important conceptual and practical issue is the degree of overlap in gene specificity between different cell types. We began by evaluating the overlap for all pairs of supercluster-level cell types. For the 435 unique supercluster pairs, the overlap was low (Jaccard index, JI, median 0.049, IQR 0.022-0.099, range 0.01-0.467). The lowest overlaps were for Amygdala excitatory-Vascular (JI = 0.01), Choroid plexus-Deep layer intratelencephalic (JI = 0.01), and Choroid plexus-Splatter (JI=0.01). The greatest overlaps were for Deep layer intratelencephalic-Upper layer intratelencephalic (JI = 0.467), Eccentric medium spiny neuron-Medium spiny neuron (JI = 0.372), and Astrocyte-Bergmann glia(JI = 0.370). As shown in *Figure S6* there were a few instances with a modest degree of clustering.

Although the overlaps were generally not marked over all pairs, all supercluster classes had at least one other class with potentially important overlap in specific genes; the maximum JI for each supercluster class had a median of 0.26 (IQR 0.19-0.33, range 0.12-0.47).

Therefore, the S-LDSC SNP-heritability enrichment of one supercluster class could be dependent on another class with overlapping TDEP genes. To examine such dependency, we performed conditional analyses in a pairwise fashion. For each supercluster class, we added the TDEP genes of another class into the S-LDSC model and evaluated how that influenced the significance of enrichment for the supercluster of interest. If the result remained significant, it means that the enrichment for the supercluster of interest is statistically independent of the effect of the supercluster being conditioned upon. For instance, the signal for the “Upper layer intratelencephalic” class became non-significant after conditioning on the “Deep layer intratelencephalic” class (*Figure 3A*), suggesting that the signal from the former was statistically dependent on the latter potentially due to overlap between TDEP genes. On the other hand, the signal of “CGE interneuron” supercluster was statistically independent from those of “MGE interneuron” and “LAMP5-LHX6 and Chandelier” despite overlap in TDEP genes (*Figure S6*).

#### Gene set analysis

Gene set analyses were conducted using hypergeometric tests versus a background of 18,090 genes. Summarizing text from the beginning of the *Methods*: the 18,090 genes are based on the GENCODE primary assembly (GRCh38.p13, v35, 3/2020, hg38) ^27,69^. In these analyses, we focused on 18,090 genes that were protein-coding, mapped to canonical autosomes (chr1 to chr22), not in the extended MHC region (chr6:25-34 mb), and expressed in ≥ 1 of the 461 cell clusters. Hypergeometric P-values were FDR-corrected. The gene set analysis is performed for the TDEP genes of the 31 superclusters, as well as the top 25 clusters for schizophrenia. The major Gene Ontology (GO) terms in the gene sets enriched for the top 25 clusters for schizophrenia were summarized in treemaps using R::rrvgo. In addition to the Cellular Component (GO-CC) category (*Figure 3C*), *Figures S7* present the Biological Processes (GO-BP) and Molecular Function (GO-MF) domains, respectively.

#### Density-based visualization of high-dimensional data

To improve understanding of these high dimensional data, we applied UMAP to visualize these data in two dimensions and HDBSCAN to identify groupings within the UMAP projection ^80,81^.

### Analysis of brain anatomic dissections

The dissection “HTHso” (supraoptic region of hypothalamus) had a high proportion of neocortical neurons, and “A35r” had a high proportion of non-neuronal cells ^27^, suggesting potential contamination or technical issues with the dissection depth or margins. We therefore excluded HTHso and A35r and focused on 104 anatomic dissections. Anatomic dissections may contain highly heterogeneous cell type compositions ^27^, and the function of the same genes can differ between cell types. Therefore, we deem that S-LDSC is most appropriate at cell type levels (i.e., superclusters and clusters) and evaluate enrichment at the level of dissections (or, detailed brain regions) by integrating the cluster-level enrichment. For visualizing cell type components in *Figure 4A*, we calculated the proportion of each cell cluster per brain dissection as the number of cells in the cluster in the dissection divided by the total number of cells in the dissection. This number was scaled into deciles for plotting. *Table S9* presents the scaled proportion and the range and mean of the actual proportion.

Of note, neuronal cell types are dominantly enriched for the SNP-heritability of psychiatric disorders, we therefore focused on the neuronal cells in calculating dissection-level enrichment. Specifically, for each anatomic dissection, the proportion of each neuronal cluster was calculated as the number of cells in the cluster in the dissection divided by the total number of neuronal cells in the dissection. Next, for each dissection, we weighted the cluster-level enrichment Z-score (from S-LDSC) of each neuronal cluster by its proportion within the dissection, and calculated the weighted sum as the Z-score of enrichment for the dissection. Finally, P-value for each dissection was calculated based on the weighted-summed Z-score.

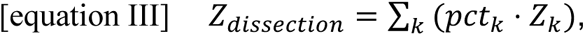

where *k* is a cluster, and 𝑝𝑐𝑡_6_ is the percentage of cluster *k* in the dissection.

### Visualizing anatomic dissection results in a 3D human brain model

The Allen Brain Atlas has a powerful 3D human brain model ^82^. As the snRNA-seq dissections were performed according to the 2D Allen Brain Atlas, we had the opportunity to transfer the labels to the 3D map, although the names of the dissections were not completely matched between the 3D and 2D Atlas. We mapped 106 snRNA-seq dissections to 84 regions in the 3D brain model ^83^ via expert curation (*Table S14*). The matched regions were used for visualizing dissection-level SNP-heritability results of schizophrenia (*Figure 4C-E*) and to obtain 3D coordinates in the fMRI analysis (*Figure 5*). Each 3D label was assigned a value as the -log_10_(P) of scz2022 SNP-heritability enrichment of the corresponding anatomic dissection in the snRNA-seq dataset. If a 3D label corresponded to multiple snRNA-seq anatomic dissections, the most significant value was taken. Not all the cerebral cortical regions in the 3D model were sampled in the snRNA-seq dataset. Given our observation that the enrichment of scz2022 heritability was generally distributed across the cerebral dissections (*Figure 4B*), we assigned the unsampled neocortical regions the mean Z-score of all the sampled neocortical regions. For unsampled regions not in the cerebral cortex, we did not make any specific assumption, and their significance of enrichment was treated as missing. ITK_SNAP (v3.8.0) was used to visualize the 3D model ^84^.

### Functional magnetic resonance imaging (fMRI)

To evaluate our findings in the context of intrinsic functional connectivity, we compared patients with schizophrenia to neurotypical controls. Resting-state fMRI scans of 46 schizophrenia cases and 46 age- and sex-matched controls from the UCLA Consortium for Neuropsychiatric Phenomics LA5c Study (https://www.openfmri.org/dataset/ds000030/) ^48^. From the public dataset, 50 cases with schizophrenia and 127 healthy controls (age: 21-50 years) were acquired. Functional scans were pre-processed, head motion-corrected, and normalized to MNI space (resolution of 3 mm^3^) ^85^. Framewise displacement (measuring the head motion during scanning) showed significantly more head motion for cases than controls. To minimize group differences, participants with framewise displacement >2mm were excluded. Next, we gradually excluded the healthy controls with fewer head motions until the difference between the groups was no longer significant. Finally, 46 patients with schizophrenia and 46 age- and sex-matched controls were included in the analysis.

In order to define an initial search space in the fMRI imaging data, we started with 45 brain regions that were mapped to the 3D brain model and showed significant schizophrenia SNP-heritability enrichment at FDR ≤ 0.01 (*Table S15*). We specified spherical regions of interest (ROIs) centering the MNI coordinates with a radius of 5mm. Regions that were too small to be identified from the fMRI data were removed, and overlapping ROIs were selected based on higher enrichment of schizophrenia SNP-heritability. Specifically, substantial overlap was found between the ROIs of the amygdala corticomedial nuclear group (CMN), namely the medial nucleus (Me), the amygdalohippocampal area (AHA), and the cortical amygdaloid nuclei (specifically the anterior part, CoA); we used the CoA (highest significance of schizophrenia SNP-heritability enrichment) to represent CMN. Likewise, the lateral nucleus (La) of the amygdala was prioritized over the nearby, overlapping structures basolateral nucleus (BL) and basomedial nucleus (BM), to represent the basolateral nuclear group (BLN). The ambiens gyrus (AG), frontal agranular insular cortex (FI), and claustrum (Cla) were too small for ROI identification and thus removed. This resulted in 38 unique brain ROIs; taking laterality into account, we considered 76 regions (38 in each hemisphere) in the following analysis. The resting state fMRI time-series of all voxels within each ROI were then averaged to obtain the regional time-series. The connectivity between two ROIs was defined as the Pearson’s correlation of the functional time-series of the two ROIs, and we fed the pairwise connectivities into a deep neural network classifier ^86^ as features to distinguish cases from controls using Braph2 ^87^.

We randomly split the sample into five equal portions (10 cases and 10 controls in portion 1, and 9 cases and 9 controls per portion in portions 2-5) and ran five folds of parallel analyses. For each fold, we took one portion as the test set and the rest as the training set, and trained a deep neural network classifier using Braph2 to distinguish cases from controls. In each fold, recursive feature elimination strategy ^88^ was adopted to reduce the number of ROIs in a stepwise manner, where the least contributing ROI was removed from the next iteration. In each run, we applied a deep neural network classifier, Braph2 (an unpublished update of Braph ^87^), to model the network to distinguish cases from controls. The deep neural network classifier was trained with five-fold cross-validation on randomly shuffled data within the training set. We applied the trained network to the independent test set in the fold and acquired the area under curve (AUC) to evaluate the network. In total, we obtained 75 data-driven networks and their AUCs per fold. The same cross validation setup was applied to the defined brain regions for the Core and Default Mode Networks, without the recursive feature elimination process^29^. Regions included in these two networks are listed in *Table S11*.

Cross-entropy loss function was used to calculate the model error in the optimization of the neural network classifier. In each run, 1000 permutations were performed to evaluate the contribution of all features (i.e. the input ROI-to-ROI pairwise connections/edges) ^89^. In this approach, a single feature value was randomly shuffled 1000 times, which in turn established the 95% confidence interval of the model error. If the model error of the original loss model fell outside the 95% confidence interval, the feature was considered as a *contributing* feature to the model. For a contributing feature, we calculated its relative feature importance (FI) as:

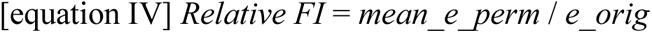

where *mean_e_perm* is the averaged model error across the permutations, and *e_orig* is the original loss without any permutation. By doing this, we obtained a FI for each feature (i.e., ROI-to-ROI pairwise connection/edge) in each cross-validation and summed those FIs across cross-validations as the *final relative FI* for the feature. Since each ROI could have several contributing features and thus several FI, we summed up all final relative FI of each ROI as the measure of the contribution of the ROI to the classifier in the specific run. The ROI with the least contribution was removed from the input for the next iteration, which realized the recursive feature elimination.

Given the variation in AUC (*Figure S8A, Table S12*), which was likely to be attributable to the limited number in each test set, we opted to describe all models with AUC>0.5 instead of picking the model with highest AUC per fold. We focused the description on two levels, namely the level of regions and the level of connections. According to recessive feature elimination described above, regions with higher contributions tend to remain during the feature reduction. We therefore counted the frequency of each region across all folds along the feature reduction runs (*Figure S8B*) and among only models with AUC>0.5 (*Figure 5B*). As expected, hippocampal and amygdalar regions indeed presented more frequently in the process, which remained true when focusing on only models with AUC>0.5. The hippocampal and subcortical regions occupied greater portions in the top observed regions (*Figure S8D*), although they were only minorities in all the included regions in the models with AUC>0.5 (*Figure S8C*). Next, at the level of connections, we counted the number of valid connections per region among all the models with AUC>0.5, where valid connections were defined as final relative FI >0.5 (“final relative FI” was introduced in the paragraph above). Next, we calculate the strength of each pairwise connection/edge/feature as below: per model per edge, weight the final relative FI by the AUC; per fold per edge, sum up the weighted final relative FI; per fold, standardize the summed weighted final relative FI for all edges to mean=0 and standard deviation=1, so the measure became comparable between folds; and finally, per edge, sum across folds the summed weighted final relative FI to obtain the final edge strength. This allowed direct comparison between the pairwise connections. The top connections were enriched in the hippocampal, subcortical, and some cortical regions (*Figure S8E, Table S13*). We plotted the distribution of these strength measures and applied a cutoff at 0.5% (*Figure S8F*), which resulted in 14 top connections to be plotted on the superior view of the brain (*Figure 5D*).

### Cell type and evolutionary constraint

As described in Sullivan et al. ^1^, we used the Zoonomia alignment of 241 placental mammals to create a gene constraint metric. In comparing multiple different constraint metrics, the simplest metric appeared to be the best (cdsFracCons, the number of constrained CDS bases divided by the total number of CDS bases). cdsFracCons does not have the limitations of alternative measures (e.g., pLI is close to a dichotomy and LOEUF has a strong residual correlation with CDS size) ^90,91^.

Code available at: https://github.com/Hjerling-Leffler-Lab/TDEP-sLDSC

---

**Figure S1.**
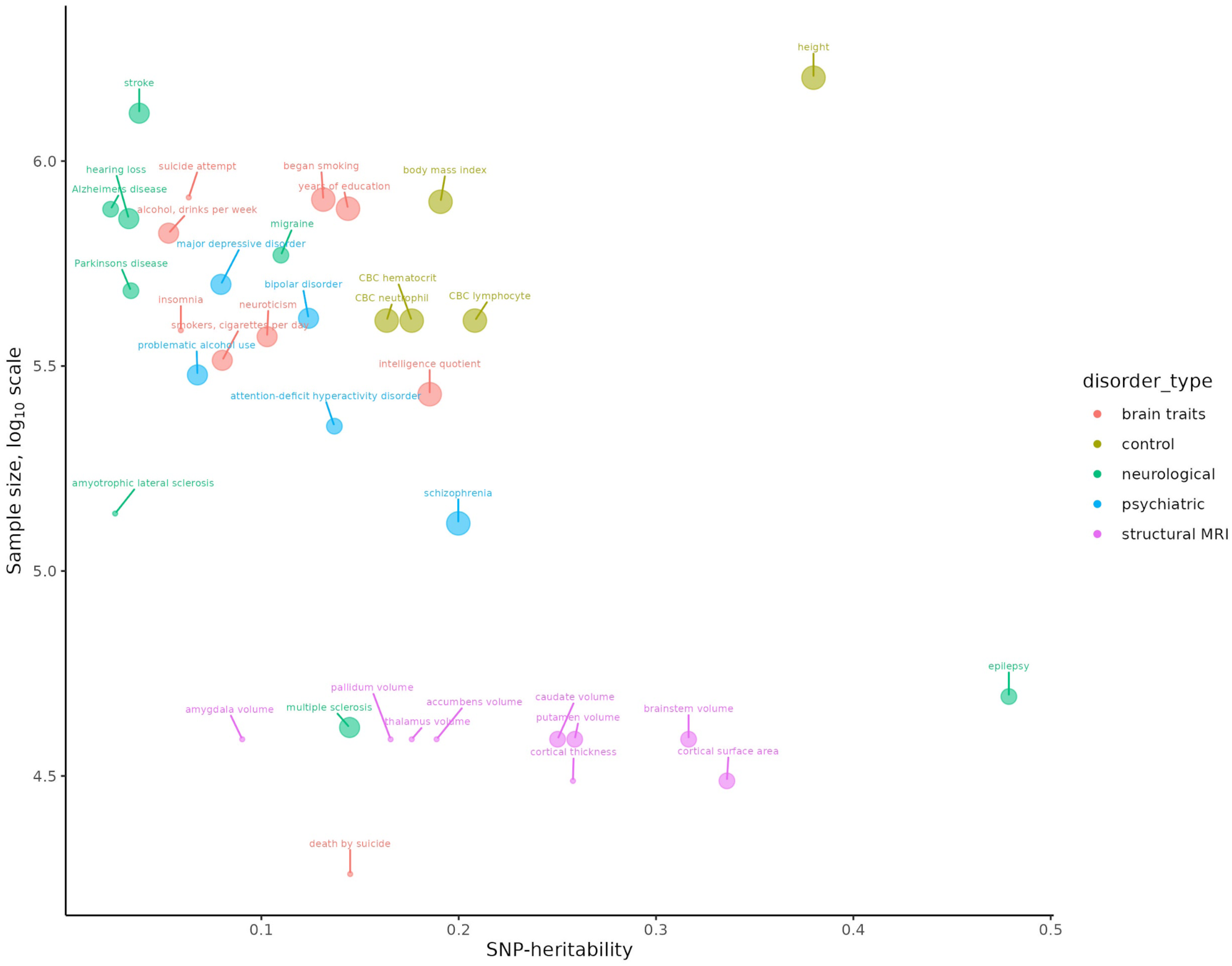
Depiction of primary GWAS trait features (data in Table S1). X-axis depicts liability-scale SNP-heritability estimated using LDSC. Y-axis is sample size on log_10_ scale. Point colors correspond to disorder type. Point size shows quartiles of the number of LD-clumped loci: quartile 1 had 3-13 loci (e.g., insomnia, suicide attempt); quartile 2 had 13-37 loci (e.g., Alzheimer’s, epilepsy); quartile 3 had 40-103 loci (e.g., bipolar disorder, MDD); and quartile 4 had 180-2705 loci (BMI, educational attainment, height).

**Figure S2.**
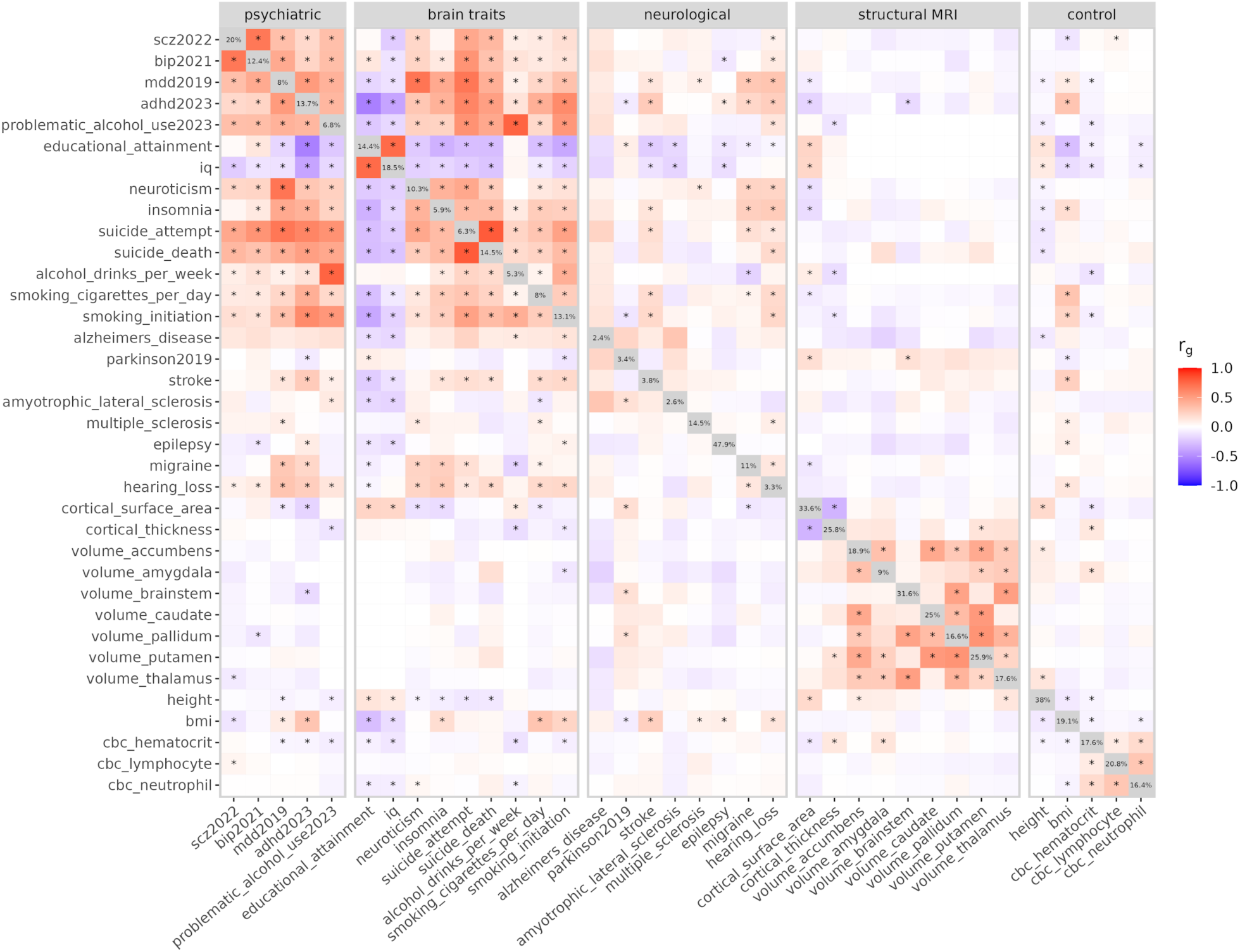
Heatmap of genetic correlations (r_g_) for the primary GWAS traits. Trait labels are defined in Table S1. We used LDSC to estimate SNP-heritability of each trait (shown on the diagonal) and genetic correlations between traits (off-diagonal values). Traits are grouped by disorder type. Colors indicate the strength and direction of genetic correlations. Asterisks indicate significant genetic correlation at FDR < 0.05 per trait. We have blanked out the genetic correlations of subcortical volumes with IQ, educational attainment, and alcohol use (the data owners prohibit use for “research into the genetics of intelligence, education, … or addictions”).

**Figure S3.**
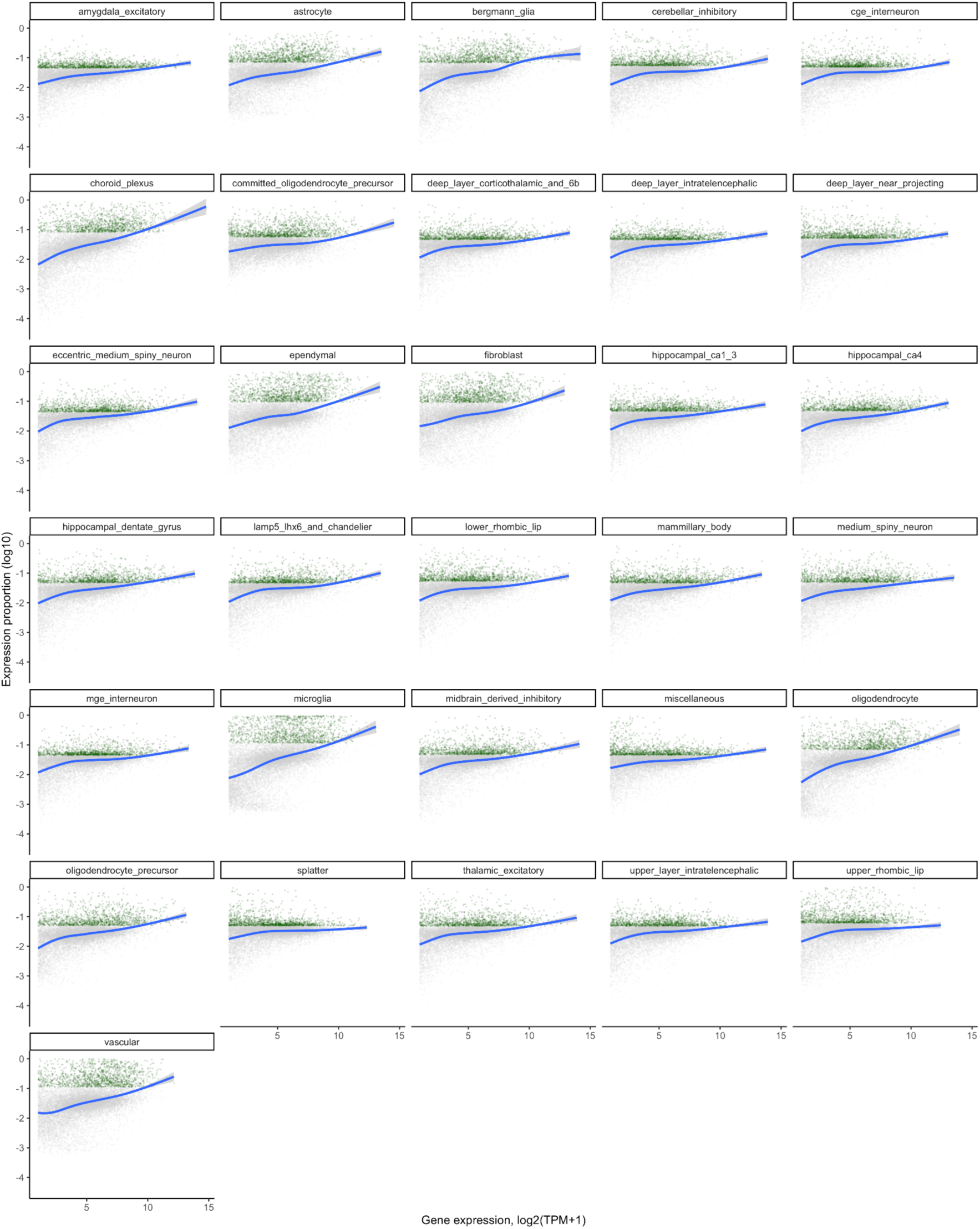
Relation of TPM (x-axis) and expression proportion (EP, y-axis) separately for each of the 31 superclusters. Both axes were log transformed to add separation. Each point is a brain-expressed protein-coding gene. Green points show the top-decile EP (TDEP) genes for a supercluster, and gray all other genes. The blue line is a lowess smoother.

**Figure S4.**
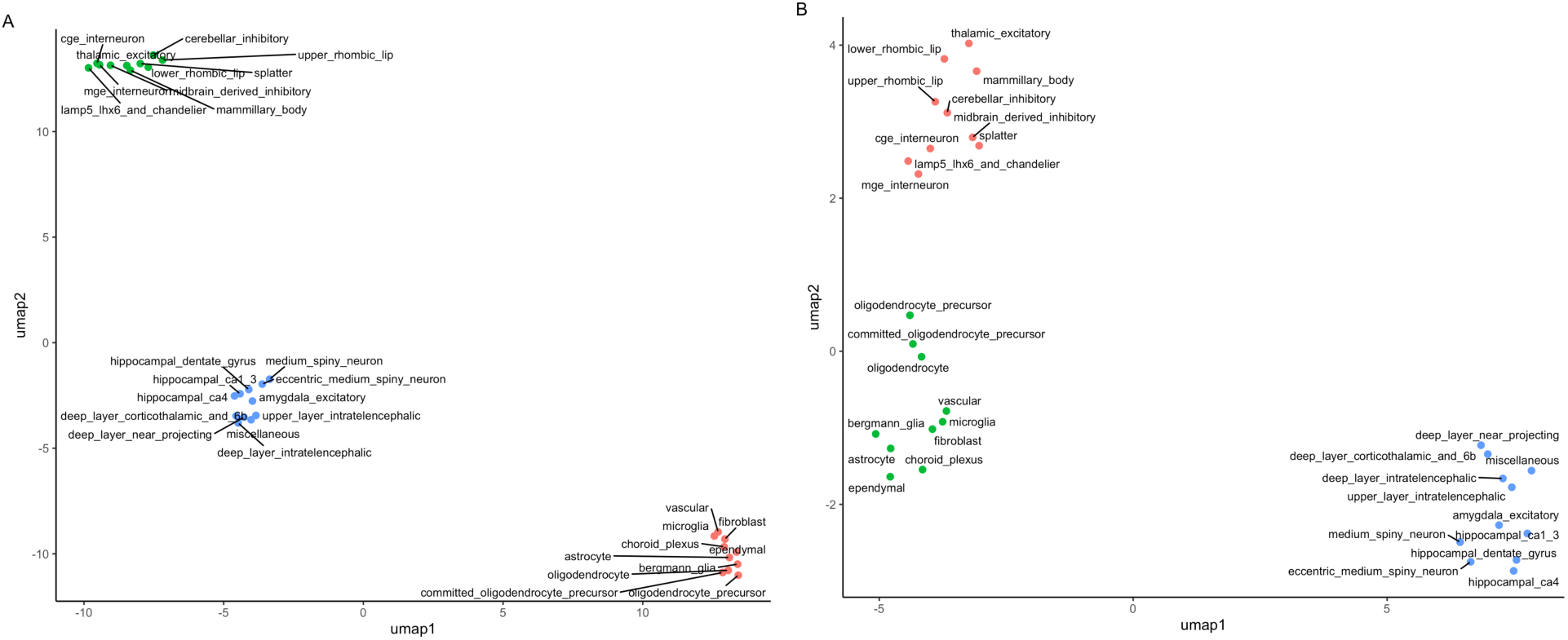
UMAP (n_neighbors=5) / HDBSCAN (minPts = 5) analysis. Panel A is for TDEP and panel B for log2(TMP+1).

**Figure S5.**
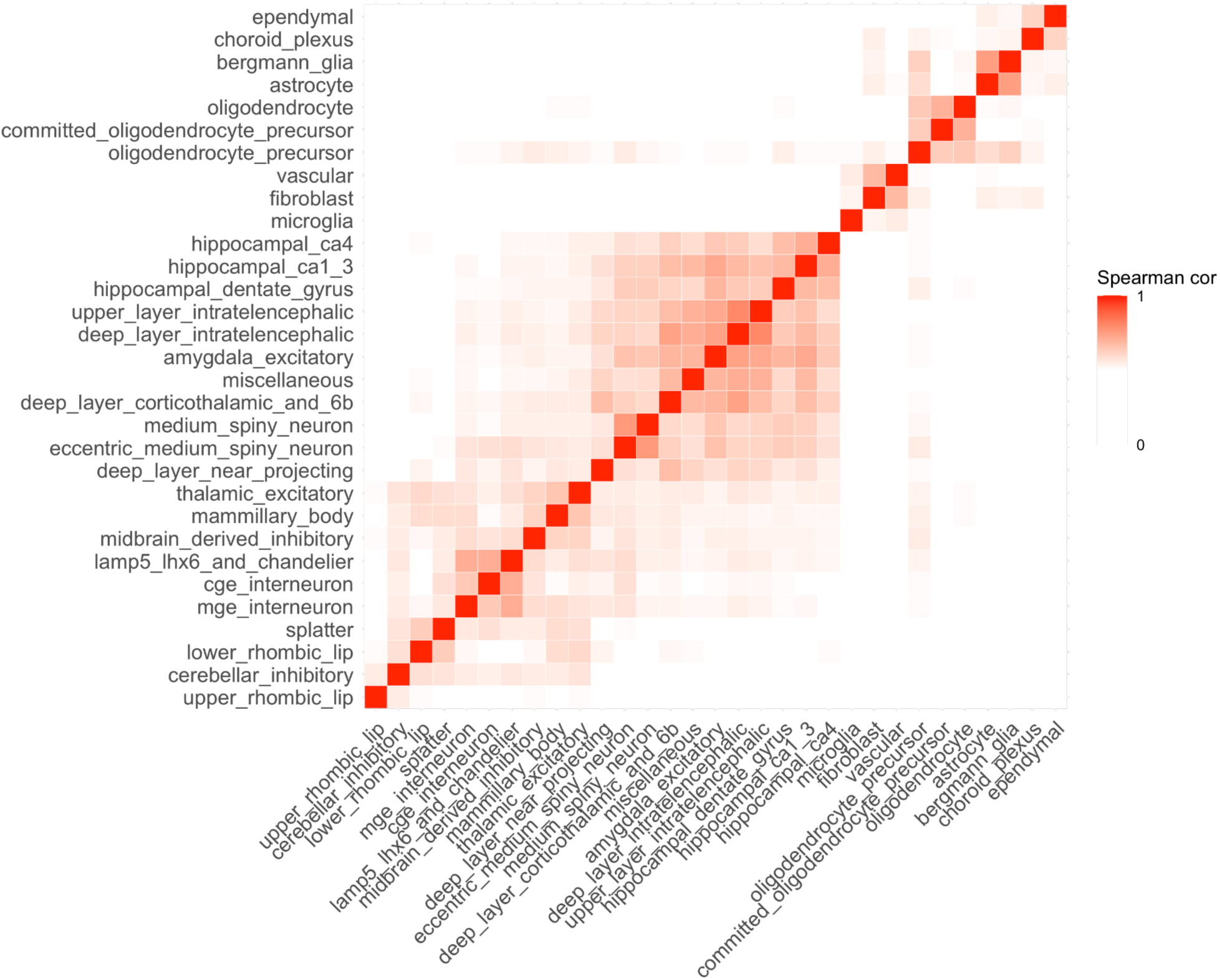
Genomic bin analysis. For each supercluster and 100 kg genomic bins, we numerically assessed the degree to which the observed number of TDEP gene TSS per bin deviated from the total number of protein-coding TSS per bin (studentized residuals). The above is a heatmap of the Spearman correlation matrix (following hierarchical clustering).

**Figure S6.**
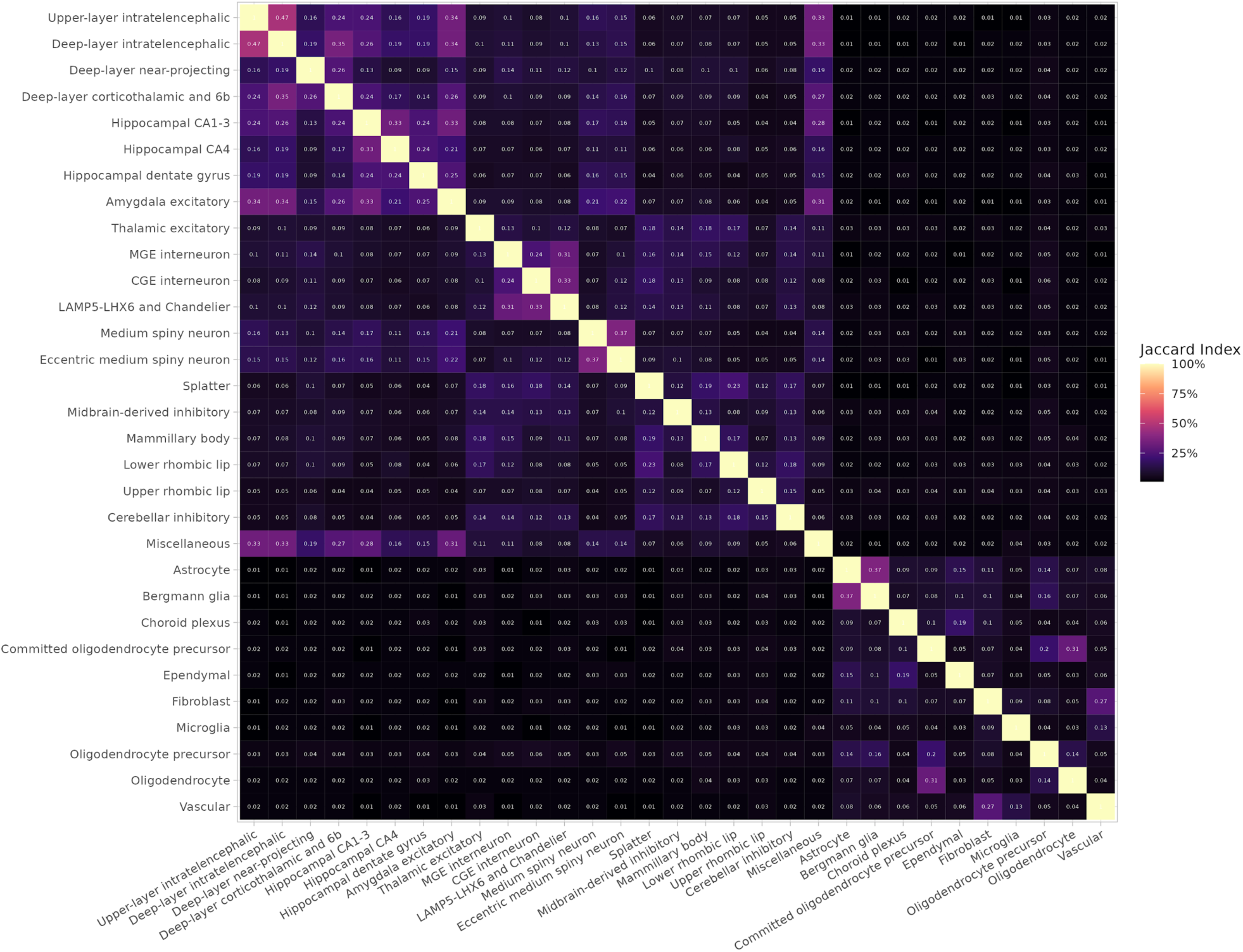
Jaccard index heatmap. For all pairs of superclusters, we computed the Jaccard index for overlap of TDEP genes (these gene sets are the input annotation for S-LDSC). For two sets of genes, Jaccard index of 1 means complete overlap and 0 means no overlap.

**Figure S7.**
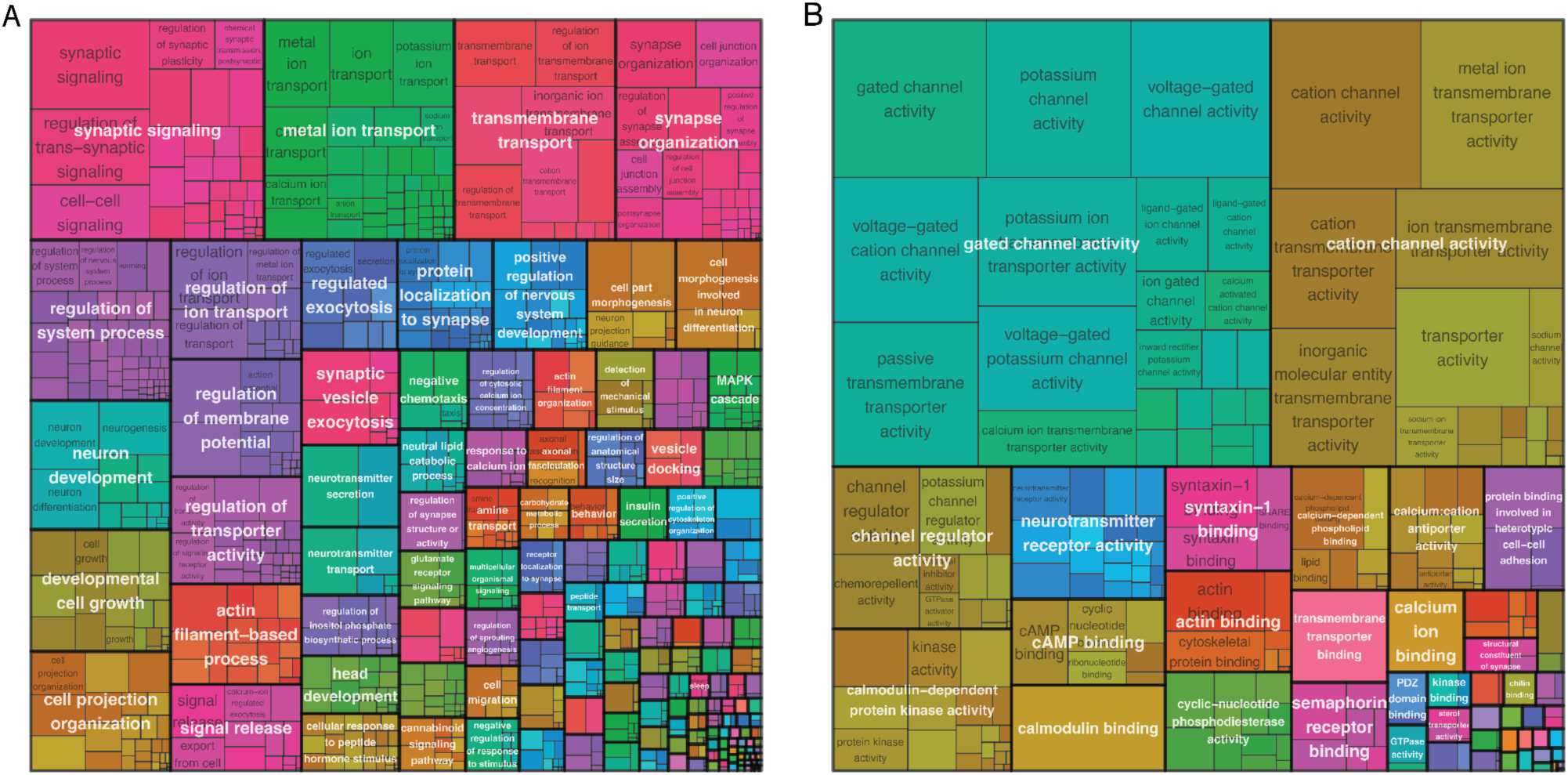
Treemap for gene ontology (GO) gene set enrichment for the TDEP genes of the top 25 clusters enriched of schizophrenia SNP-heritability. (A) GO-BP gene sets enriched in the clusters. (B) GO-MF gene sets enriched in the clusters.

**Figure S8.**
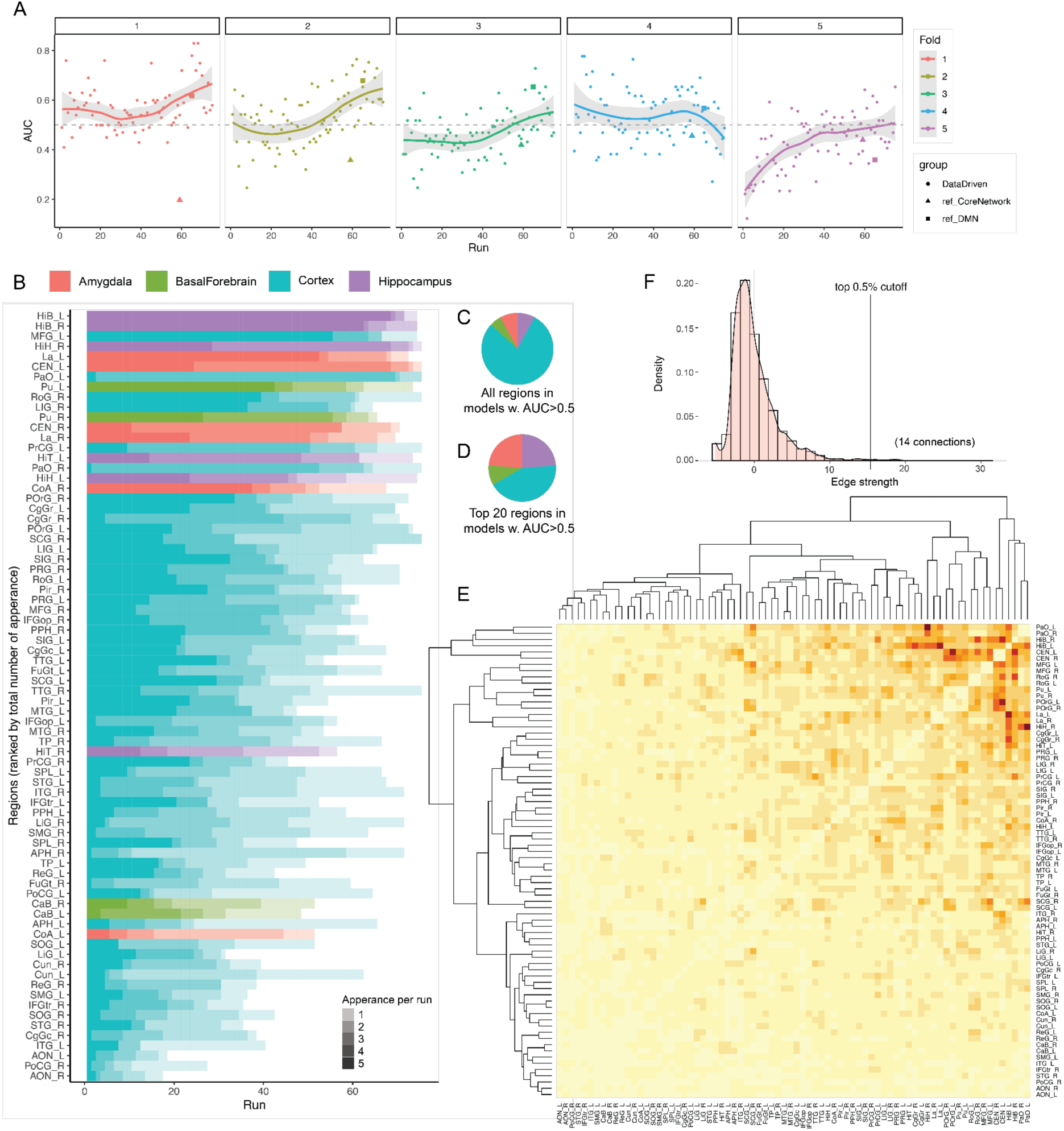
fMRI analysis supplementary figures. (A) AUC of each model across the 5 parallel folds. Round dots are AUCs of the data-driven networks/models. Triangle dots are the AUCs of the core network, and the square dots are the AUCs of the default mode network (DMN); these two reference networks were included once each in every fold and they were plotted at the Run (x-axis) that corresponds to their sizes. (B) Regions ranked by the total number of appearances across all 5 folds in each run. The shade indicates the number of appearances, and the color indicates the broader brain areas. (C)-(D): pie-charts showing the proportion of each broader brain area in all regions in models with AUC>0.5 (C) and in the top 20 regions in these models (D), which corresponds to the bar plot in Figure 5B. Although hippocampal and amygdalar regions were relatively small portions of all the regions, they occupied larger portions among the top important regions. (E) Heatmap of the pairwise connections included in all models with AUC>0.5. The strength of each connection/edge was calculated as below: per model per edge, weight the final relative FI by the AUC; per fold per edge, sum up the weighted final relative FI; per fold, standardize the summed weighted final relative FI for all edges to mean=0 and standard deviation=1, so the measures are comparable between folds; and finally, per edge, sum across folds the summed weighted final relative FI to obtain the final edge strength. (F) The density plot of the final edge strength for all the edges/connections/features in panel (E). We applied a cutoff of the top 0.5% to select the most important edges/connections/features for plotting in Figure 5D.

**Table S1.** Summary of GWAS included. The image shows 10 rows. The full table is in the supplemental-tables file.

| primary_gwas | disorder_type | phenotype | label | year | pmid | trait_type | n | ncas | ncon | nsig_snps_postQC | n_loci | lambda_gc | ldsc_intercept | ldsc_h2 |
| --- | --- | --- | --- | --- | --- | --- | --- | --- | --- | --- | --- | --- | --- | --- |
| TRUE | psychiatric | schizophrenia | scz2022 | 2022 | 35396580 | binary | 130644 | 53386 | 77258 | 17420 | 180 | 1.777 | 1.101 | 0.203 |
| TRUE | psychiatric | bipolar disorder | bip2021 | 2021 | 34002096 | binary | 413466 | 41917 | 371549 | 2702 | 57 | 1.471 | 1.038 | 0.126 |
| TRUE | psychiatric | major depressive disorder | mdd2019 | 2019 | 30718901 | binary | 500199 | 170756 | 329443 | 3883 | 51 | 1.453 | 1.002 | 0.080 |
| TRUE | psychiatric | attention-deficit hyperactivity disorder | adhd2023 | 2023 | 36702997 | binary | 225534 | 38691 | 186843 | 1213 | 27 | 1.362 | 1.043 | 0.137 |
| TRUE | psychiatric | problematic alcohol use | problematic_alcohol_use2023 | 2023 | 32451486 | combined | 300789 |  |  | 4124 | 81 | 1.573 | 1.039 | 0.068 |
| TRUE | brain traits | years of education | educational_attainment | 2022 | 35361970 | continuous | 765283 |  |  | 58790 | 906 | 2.593 | 1.219 | 0.145 |
| TRUE | brain traits | intelligence quotient | iq | 2018 | 29942086 | continuous | 269867 |  |  | 10875 | 210 | 1.922 | 1.083 | 0.186 |
| TRUE | brain traits | neuroticism | neuroticism | 2018 | 29942085 | continuous | 372903 |  |  | 6591 | 103 | 1.607 | 1.030 | 0.104 |
| TRUE | brain traits | insomnia | insomnia | 2019 | 30804565 | binary | 386533 | 109402 | 277131 | 393 | 13 | 1.310 | 1.015 | 0.060 |

## Data Availability

All data produced in the present study are available upon reasonable request to the authors

https://github.com/Hjerling-Leffler-Lab/TDEP-sLDSC

## Acknowledgements

JHL was supported by the Swedish Research Council (Vetenskapsrådet, award 2018-00799), Swedish Brain Foundation (Hjärnfonden, award FO2018-0272) and European Research Council (SCHIZTYPE, grant agreement 819540). PFS was supported by the Swedish Research Council (Vetenskapsrådet, award D0886501) and the US National Institute of Mental Health (R01s MH124871, MH121545, and MH123724). YL was supported by the European Research Council (SUBTREAT, grant agreement 101042183) and the US National Institute of Mental Health (R01 MH123724). SY was supported by the StratNeuro postdoctoral grant.

We would like to acknowledge the International Headache Genetics Consortium for sharing the GWAS summary statistics on migraine.

## Collaborators

### International Suicide Genetics Consortium

Anna R Docherty^1,2,3^, Niamh Mullins^4,5^, Allison E Ashley-Koch^6^, Xuejun Qin^6^, Jonathan R I Coleman^7,8^, Andrey Shabalin^1,2^, JooEun Kang^9^, Balasz Murnyak^1,2^, Frank Wendt^10^, Mark Adams^11^, Adrian I ^Campos12,13^, Emily DiBlasi^1,2^, Janice M Fullerton^14,15^, Henry R Kranzler^16,17^, Amanda Bakian^2^, Eric T Monson^2^, Miguel E Rentería^12,18^, Consuelo Walss-Bass^19^, Ole A Andreassen^20,21^, Cynthia M Bulik^22,23,24^, Howard J Edenberg^25,26^, Ronald C Kessler^27^, J John Mann^28^, John I Nurnberger Jr^29,29^, Giorgio Pistis^30^, Fabian Streit^31^, Robert J Ursano^32^, Renato Polimonti^10^, Michelle Dennis^33^, Melanie Garrett^34^, Lauren Hair^35^, Philip Harvey^36^, Elizabeth R Hauser^6,37^, Michael A Hauser^6^, Jennifer Huffman^38^, Daniel Jacobson^39^, Jennifer H Lindquist^40^, Ravi Madduri^41^, Benjamin McMahon^42^, David W Oslin^43,44^, Jodie Trafton^45^, Swapnil Awasthi^46^, Andrew W Bergen^47,48^, Wade H Berrettini^49^, Martin Bohus^50^, Harry Brandt^51,52^, Xiao Chang^53^, Hsi-Chung Chen^54^, Wei J Chen^54,55,56^, Erik D Christensen^57,58^, Steven Crawford^51,52^, Scott Crow^59^, Philibert Duriez^60,61^, Alexis C Edwards^3^, Fernando Fernández-Aranda^62^, Manfred M Fichter^63,64^, Hanga Galfalvy^65,66^, Steven Gallinger^67^, Michael Gandal^68^, Philip Gorwood^60,61^, Yiran Guo^53^, Jonathan D Hafferty^11^, Hakon Hakonarson^53,69^, Katherine A Halmi^70^, Akitoyo Hishimoto^71^, Sonia Jain^72^, Stéphane Jamain^73^, Susana Jiménez-Murcia^62^, Craig Johnson^74^, Allan S Kaplan^75,76,77^, Walter H Kaye^78^, Pamela K Keel^79^, James L Kennedy^75,76,77^, Minsoo Kim^68^, Kelly L Klump^80^, Daniel F Levey^81,82^, Dong Li^53^, Shih-Cheng Liao^54^, Klaus Lieb^83^, Lisa Lilenfeld^84^, Adriana Lori^85^, Pierre J Magistretti^86,87^, Christian R Marshall^88^, James E Mitchell^89^, Richard M Myers^90^, Satoshi Okazaki^91^, Ikuo Otsuka^66,91^, Dalila Pinto^4,5^, Abigail Powers^85^, Nicolas Ramoz^61^, Stephan Ripke^46,92,93^, Stefan Roepke^94^, Vsevolod Rozanov^95,96^, Stephen W Scherer^97,98^, Christian Schmahl^50^, Marcus Sokolowski^99^, Anna Starnawska^100,101,102,103^, Michael Strober^104,105^, Mei-Hsin Su^56^, Laura M Thornton^24^, Janet Treasure^106,107^, Erin B Ware^108,109^, Hunna J Watson^24,110,111^, Stephanie H Witt^31^, D Blake Woodside^76,77,112,113^, Zeynep Yilmaz^24,114,115^, Lea Zillich^31^, Rolf Adolfsson^116^, Ingrid Agartz^117,118,119^, Tracy M Air^120^, Martin Alda^121,122^, Lars Alfredsson^123,124^, Adebayo Anjorin^125^, Vivek Appadurai^126,127^, María Soler Artigas^128,129,130,131^, Sandra Van der Auwera^132,133^, M Helena Azevedo^134^, Nicholas Bass^135^, Claiton HD Bau^136,137^, Bernhard T Baune^138,139^, Frank Bellivier^140,141,142,143^, Klaus Berger^144^, Joanna M Biernacka^145^, Tim B Bigdeli^3,146^, Elisabeth B Binder^85,147^, Michael Boehnke^148^, Marco P Boks^149^, Rosa Bosch^128,129,150^, David L Braff^151^, Richard Bryant^152^, Monika Budde^153^, Enda M Byrne^13,154^, Wiepke Cahn^155^, Miguel Casas^128,129,131,150^, Enrique Castelao^30^, Jorge A Cervilla^156^, Boris Chaumette^157,158,159^, Sven Cichon^160,161,162,163^, Aiden Corvin^164^, Nicholas Craddock^165^, David Craig^166^, Franziska Degenhardt^163^, Srdjan Djurovic^167,168^, Ayman H Fanous^3,146^, Jerome C Foo^169^, Andreas J Forstner^160,163,170^, Mark Frye^171^, Justine M Gatt^14,152^, Pablo V Gejman^172,173^, Ina Giegling^174,175^, Hans J Grabe^132,133^, Melissa J Green^14,176^, Eugenio H Grevet^177,178^, Maria Grigoroiu-Serbanescu^179^, Blanca Gutierrez^180^, Jose Guzman-Parra^181^, Steven P Hamilton^182^, Marian L Hamshere^165^, Annette M Hartmann^174^, Joanna Hauser^183^, Stefanie Heilmann-Heimbach^163^, Per Hoffmann^161,162,163^, Marcus Ising^184^, Ian Jones^165^, Lisa A Jones^185^, Lina Jonsson^186^, René S Kahn^5,187^, John R Kelsoe^151,188^, Kenneth S Kendler^3^, Stefan Kloiber^75,184,189^, Karestan C Koenen^92,190,191^, Manolis Kogevinas^192^, Bettina Konte^174^, Marie-Odile Krebs^157,158,159^, Mikael Landén^22,193^, Jacob Lawrence^194^, Marion Leboyer^195,196,197^, Phil H Lee^92,93,198^, Douglas F Levinson^199^, Calwing Liao^200,201^, Jolanta Lissowska^202^, Susanne Lucae^184^, Fermin Mayoral^181^, Susan L McElroy^203^, Patrick McGrath^204^, Peter McGuffin^8^, Andrew McQuillin^135^, Divya Mehta^205,206^, Ingrid Melle^20,207^, Yuri Milaneschi^208^, Philip B Mitchell^176^, Esther Molina^209^, Gunnar Morken^210,211^, Preben Bo Mortensen^101,114,127,212^, Bertram Müller-Myhsok^147,213,214^, Caroline Nievergelt^151^, Vishwajit Nimgaonkar^215^, Markus M Nöthen^163^, Michael C O’Donovan^165^, Roel A Ophoff^68,216^, Michael J Owen^165^, Carlos Pato^217,217^, Michele T Pato^218^, Brenda WJH Penninx^219^, Jonathan Pimm^135^, James B Potash^220^, Robert A Power^8,221,222^, Martin Preisig^30^, Digby Quested^223^, Josep Antoni Ramos-Quiroga^128,129,131,150^, Andreas Reif^224^, Marta Ribasés^128,129,130,131^, Vanesa Richarte^128,129,150^, Marcella Rietschel^225^, Margarita Rivera^8,226^, Andrea Roberts^227^, Gloria Roberts^176^, Guy A Rouleau^228,229^, Diego L Rovaris^230^, Dan Rujescu^174^, Cristina Sánchez-Mora^128,129,130,131^, Alan R Sanders^172,173^, Peter R Schofield^14,15^, Thomas G Schulze^153,169,231,232,233^, Laura J Scott^148^, Alessandro Serretti^234^, Jianxin Shi^235^, Stanley I Shyn^236^, Lea Sirignano^169^, Pamela Sklar^4,5,237^, Olav B Smeland^20,21^, Jordan W Smoller^92,191,238^, Edmund J S Sonuga-Barke^239^, Gianfranco Spalletta^240,241^, John S Strauss^75,189^, Beata Świątkowska^242^, Maciej Trzaskowski^13^, Ming T Tsuang^243^, Gustavo Turecki^244^, Laura Vilar-Ribó^128,131^, John B Vincent^245^, Henry Völzke^246^, James TR Walters^165^, Cynthia Shannon Weickert^14,176^, Thomas W Weickert^14,176^, Myrna M Weissman^247,248^, Leanne M Williams^249^, Naomi R Wray^13,206^, Clement C Za_i_^92,189,190,250,251,252^, Esben Agerbo^114,212,253^, Anders D Børglum^100,101,102,103^, Gerome Breen^7,8^, Ditte Demontis^100,101,102,103^, Annette Erlangsen^103,254,255,256^, Tõnu Esko^257,258^, Joel Gelernter^81,82^, Stephen J Glatt^259^, David M Hougaard^253,260^, Hai-Gwo Hwu^261^, Po-Hsiu Kuo^54,56^, Cathryn M Lewis^8,262^, Qingqin S Li^263^, Chih-Min Liu^54^, Nicholas G Martin^12^, Andrew M McIntosh^11^, Sarah E Medland^12^, Ole Mors^253,264^, Merete Nordentoft^253,265^, Catherine M Olsen^266^, David Porteous^267^, Daniel J Smith^268^, Eli A Stahl^4,257,269^, Murray B Stein^270^, Danuta Wasserman^99^, Thomas Werge^126,253,271,272^, David C Whiteman^266^, Virginia Willour^273^, the VA Million Veteran Program (MVP), the MVP Suicide Exemplar Workgroup, Suicide Working Group of the Psychiatric Genomics Consortium, Major Depressive Disorder Working Group of the Psychiatric Genomics Consortium, Bipolar Disorder Working Group of the Psychiatric Genomics Consortium, Schizophrenia Working Group of the Psychiatric Genomics Consortium, Eating Disorder Working Group of the Psychiatric Genomics Consortium, German Borderline Genomics Consortium, Hilary Coon^1,2,274^, Jean C Beckham^275,276^, Nathan A Kimbrel^275,276^, Douglas M Ruderfer^9,277,278^

^1^ Huntsman Mental Health Institute, Salt Lake City, UT, USA

^2^ University of Utah School of Medicine, Department of Psychiatry, Salt Lake City, UT, USA

^3^ Virginia Commonwealth University, Department of Psychiatry, Richmond, VA, USA

^4^ Icahn School of Medicine at Mount Sinai, Department of Genetics and Genomic Sciences, New York, NY, USA

^5^ Icahn School of Medicine at Mount Sinai, Department of Psychiatry, New York, NY, USA

^6^ Duke University Medical Center, Duke Molecular Physiology Institute, Durham, NC, USA

^7^ King’s College London, National Institute for Health Research (NIHR) Maudsley Biomedical Research Centre at South London and Maudsley NHS Foundation Trust, London, UK

^8^ King’s College London, Social Genetic and Developmental Psychiatry Centre, London, UK

^9^ Vanderbilt University Medical Center, Division of Genetic Medicine, Department of Medicine, Vanderbilt Genetics Institute, Nashville, TN, USA

^10^ Yale University School of Medicine, Department of Psychiatry, New Haven, CT, USA

^11^ University of Edinburgh, Division of Psychiatry, Edinburgh, UK

^12^ QIMR Berghofer Medical Research Institute, Mental Health and Neuroscience Research Program, Brisbane, QLD, Australia

^13^ The University of Queensland, Institute for Molecular Bioscience, Brisbane, QLD, Australia

^14^ Neuroscience Research Australia, Sydney, NSW, Australia

^15^ University of New South Wales, School of Medical Sciences, Sydney, NSW, Australia

^16^ University of Pennsylvania Perelman School of Medicine, Department of Psychiatry, Philadelphia, PA, USA

^17^ Crescenz VAMC, VISN 4 MIRECC, Philadelphia, PA, USA

^18^ The University of Queensland, School of Biomedical Sciences, Faculty of Medicine, Brisbane, QLD, Australia

^19^ University of Texas Health Science Center, Department of Psychiatry and Behavioral Sciences, Houston, TX, USA

^20^ Oslo University Hospital, Division of Mental Health and Addiction, Oslo, Norway

^21^ University of Oslo, NORMENT, Oslo, Norway

^22^ Karolinska Institutet, Department of Medical Epidemiology and Biostatistics, Stockholm, Sweden

^23^ University of North Carolina at Chapel Hill, Department of Nutrition, Chapel Hill, NC, USA

^24^ University of North Carolina at Chapel Hill, Department of Psychiatry, Chapel Hill, NC, USA

^25^ Indiana University, Department of Medical & Molecular Genetics, Indianapolis, IN, USA

^26^ Indiana University School of Medicine, Biochemistry and Molecular Biology, Indianapolis, IN, USA

^27^ Harvard Medical School, Department of Health Care Policy, Boston, MA, USA

^28^ Columbia University, Departments of Psychiatry and Radiology, New York, NY, USA

^29^ Indiana University School of Medicine, Departments of Psychiatry and Medical and Molecular Genetics, Indianapolis, IN, USA

^30^ Lausanne University Hospital and University of Lausanne, Department of Psychiatry, Lausanne, Vaud, Switzerland

^31^ Central Institute of Mental Health, Medical Faculty Mannheim, University of Heidelberg, Department of Genetic Epidemiology in Psychiatry, Mannheim, Germany

^32^ Uniformed Services University of the Health Sciences, Department of Psychiatry, Bethesda, MD, USA

^33^ Duke University Medical Center, Department of Psychiatry and Behavioral Sciences, Durham, NC, USA

^34^ Duke University Medical Center, Durham, NC, USA

^35^ Durham Veterans Affairs Health Care System, Durham, NC, USA

^36^ Miami VA Health Care System, Miami, FL, USA

^37^ Durham Veterans Affairs Health Care System, Cooperative Studies Program Epidemiology Center, Durham, NC, USA

^38^ Boston VA Health Care System, Boston, MA, USA

^39^ Oak Ridge National Laboratory, Oak Ridge, TN, USA

^40^ Durham Veterans Affairs Health Care System, VA Health Services Research and Development Center of Innovation to Accelerate Discovery and Practice Transformation, Durham, NC, USA

^41^ Argonne National Laboratory, University of Chicago Consortium for Advanced Science and Engineering, Chicago, IL, USA

^42^ Los Alamos National Laboratory, Theoretical Division, Los Alamos National Laboratory, Los Alamos, NM, USA

^43^ Corporal Michael J. Crescenz VA Medical Center, VISN 4 Mental Illness Research, Education, and Clinical Center, Philadelphia, PA, USA

^44^ Perelman School of Medicine, University of Pennsylvania, Department of Psychiatry, Philadelphia, PA, USA

^45^ VA Palo Alto Health Care System, VA Program Evaluation and Resource Center, Palo Alto, CA, USA

^46^ Charité - Universitätsmedizin Berlin, Department of Psychiatry and Psychotherapy, Berlin, Germany

^47^ BioRealm, LLC, Walnut, CA, USA

^48^ Oregon Research Institute, Eugene, OR, USA

^49^ Perelman School of Medicine at the University of Pennsylvania, Department of Psychiatry, Center for Neurobiology and Behavior, Philadelphia, PA, USA

^50^ Central Institute of Mental Health, Medical Faculty Mannheim, University of Heidelberg, Department of Psychosomatic Medicine and Psychotherapy, Mannheim, Germany

^51^ ERCPathlight, Baltimore, MD, USA

^52^ University of Maryland St. Joseph Medical Center, Baltimore, MD, USA

^53^ Children’s Hospital of Philadelphia, Center for Applied Genomics, Philadelphia, PA, USA

^54^ National Taiwan University Hospital, Department of Psychiatry, Taipei, Taiwan

^55^ National Health Research Institutes, Center for Neuropsychiatric Research, Miaoli County, Taiwan

^56^ National Taiwan University, Institute of Epidemiology and Preventive Medicine, College of Public Health, Taipei, Taiwan

^57^ Utah Department of Health and Human Services, Utah Office of the Medical Examiner, Taylorsville, UT, USA

^58^ University of Utah, Department of Pathology, Salt Lake City, UT, USA

^59^ University of Minnesota, Department of Psychiatry, Minneapolis, MN, USA

^60^ GHU Paris Psychiatrie et Neurosciences, Hôpital Sainte Anne, Paris, France

^61^ Université Paris Cité, Institute of Psychiatry and Neuroscience of Paris (IPNP), INSERM U1266, Paris, France

^62^ University Hospital Bellvitge-IDIBELL and CIBEROBN, Department of Psychiatry, Barcelona, Spain

^63^ Ludwig-Maximilians-University (LMU), Department of Psychiatry and Psychotherapy, Munich, Germany

^64^ Schön Klinik Roseneck affiliated with the Medical Faculty of the University of Munich (LMU), Munich, Germany

^65^ Columbia University, Department of Biostatistics, New York, NY, USA

^66^ Columbia University, Department of Psychiatry, New York, NY, USA

^67^ University of Toronto, Department of Surgery, Faculty of Medicine, Toronto, Canada

^68^ University of California, Los Angeles, Department of Psychiatry and Biobehavioral Science, Semel Institute, David Geffen School of Medicine, Los Angeles, CA, USA

^69^ University of Pennsylvania, The Perelman School of Medicine, Philadelphia, PA, USA

^70^ Weill Cornell Medical College, Department of Psychiatry, New York, NY, USA

^71^ Yokohama City University Graduate School of Medicine, Department of Psychiatry, Yokohama, Japan

^72^ University of California San Diego, Biostatistics Research Center, Herbert Wertheim School of Public Health and Human Longevity Science, La Jolla, CA, USA

^73^ Univ Paris-Est-Créteil, INSERM, IMRB, Translational Neuropsychiatry, Fondation FondaMental, Créteil, France

^74^ Eating Recovery Center, Denver, CO, USA

^75^ Centre for Addiction and Mental Health, Toronto, ON, Canada

^76^ University of Toronto, Department of Psychiatry, Toronto, Canada

^77^ University of Toronto, Institute of Medical Science, Toronto, Canada

^78^ University of California San Diego, Department of Psychiatry, San Diego, CA, USA

^79^ Florida State University, Department of Psychology, Tallahassee, FL, USA

^80^ Michigan State University, Department of Psychology, Lansing, MI, USA

^81^ Veterans Affairs Connecticut Healthcare Center, Department of Psychiatry, West Haven, CT, USA

^82^ Yale University School of Medicine, Division of Human Genetics, Department of Psychiatry, New Haven, CT, USA

^83^ University Medical Center, Department of Psychiatry and Psychotherapy, Mainz, Germany

^84^ The Chicago School of Professional Psychology, Washington DC, Department of Clinical Psychology, Washington, DC, USA

^85^ Emory University School of Medicine, Department of Psychiatry and Behavioral Sciences, Atlanta, GA, USA

^86^ King Abdullah University of Science and Technology, BESE Division, Thuwal, Saudi Arabia

^87^ University of Lausanne-University Hospital of Lausanne (UNIL-CHUV), Department of Psychiatry, Lausanne, Switzerland

^88^ The Hospital for Sick Children, Department of Paediatric Laboratory Medicine, Toronto, Canada

^89^ University of North Dakota School of Medicine and Health Sciences, Department of Psychiatry and Behavioral Science, Fargo, ND, USA

^90^ HudsonAlpha Institute for Biotechnology, Huntsville, AL, USA

^91^ Kobe University Graduate School of Medicine, Department of Psychiatry, Kobe, Japan

^92^ Broad Institute, Stanley Center for Psychiatric Research, Cambridge, MA, USA

^93^ Massachusetts General Hospital, Analytical and Translational Genetics Unit, Boston, MA, USA

^94^ Charité - Universitätsmedizin Berlin, Corporate Member of Freie Universität Berlin, Humboldt-Universität zu Berlin, Berlin Institute of Health, Campus Benjamin Franklin, Department of Psychiatry, Berlin, Germany

^95^ Saint-Petersburg State University, Department of Psychology, Saint-Petersburg, Russian Federation

^96^ V.M. Bekhterev National Medical Research Center for Psychiatry and Neurology, Department of Borderline Disorders and Psychotherapy, Saint-Petersburg, Russian Federation

^97^ The Hospital for Sick Children, Department of Genetics and Genomic Biology, Toronto, Canada

^98^ University of Toronto, McLaughlin Center, Toronto, Canada

^99^ Karolinska Institutet, National Centre for Suicide Research and Prevention of Mental Ill-Health (NASP), LIME, Stockholm, Sweden

^100^ Aarhus University, Centre for Genomics and Personalized Medicine, CGPM, Aarhus, Denmark

^101^ Aarhus University, Centre for Integrative Sequencing, iSEQ, Aarhus, Denmark

^102^ Aarhus University, Department of Biomedicine, Aarhus, Denmark

^103^ Aarhus University, The Lundbeck Foundation Initiative for Integrative Psychiatric Research, iPSYCH, Aarhus, Denmark

^104^ University of California Los Angeles, David Geffen School of Medicine, Los Angeles, LA, USA

^105^ University of California Los Angeles, Department of Psychiatry and Biobehavioral Science, Semel Institute for Neuroscience and Human Behavior, Los Angeles, LA, USA

^106^ King’s College London, Institute of Psychiatry, Psychology and Neuroscience, Department of Psychological Medicine, London, UK

^107^ King’s College London and South London and Maudsley National Health Service Foundation Trust, National Institute for Health Research Biomedical Research Centre, London, UK

^108^ University of Michigan, Population Studies Center, Institute for Social Research, Ann Arbor, MI, USA

^109^ University of Michigan, Survery Research Center, Institute for Social Research, Ann Arbor, MI, USA

^110^ Curtin University, School of Psychology, Perth, Western Australia, Australia

^111^ The University of Western Australia, Division of Paediatrics, Perth, Western Australia, Australia

^112^ University Health Network, Centre for Mental Health, Toronto, Canada

^113^ University Health Network, Program for Eating Disorders, Toronto, Canada

^114^ Aarhus University, National Centre for Register-Based Research, Aarhus, Denmark

^115^ University of North Carolina at Chapel Hill, Department of Genetics, Chapel Hill, NC, USA

^116^ Umeå University Medical Faculty, Department of Clinical Sciences, Psychiatry, Umeå, Sweden

^117^ Diakonhjemmet Hospital, Department of Psychiatric Research, Oslo, Norway

^118^ Karolinska Institutet, Department of Clinical Neuroscience, Centre for Psychiatry Research, Stockholm, Sweden

^119^ University of Oslo, NORMENT, Institute of Clinical Medicine, Oslo, Norway

^120^ University of Adelaide, Discipline of Psychiatry, Adelaide, SA, Australia

^121^ Dalhousie University, Department of Psychiatry, Halifax, NS, Canada

^122^ National Institute of Mental Health, Klecany, CZ

^123^ Karolinska Institutet, Department of Clinical Neuroscience, Stockholm, Sweden

^124^ Karolinska Institutet, Inst of Environmental Medicine, Stockholm, Sweden

^125^ Berkshire Healthcare NHS Foundation Trust, Psychiatry, Bracknell, UK

^126^ Copenhagen University Hospital, Institute of Biological Psychiatry, Copenhagen Mental Health Services, Copenhagen, Denmark

^127^ iPSYCH, The Lundbeck Foundation Initiative for Integrative Psychiatric Research, Copenhagen, Denmark

^128^ Hospital Universitari Vall d’Hebron, Department of Psychiatry, Barcelona, Spain

^129^ Instituto de Salud Carlos III, Biomedical Network Research Centre on Mental Health (CIBERSAM), Madrid, Spain

^130^ University of Barcelona, Department of Genetics, Microbiology & Statistics, Barcelona, Spain

^131^ Vall d’Hebron Research Institute (VHIR), Universitat Autònoma de Barcelona, Psychiatric Genetics Unit, Group of Psychiatry, Mental Health and Addiction,, Barcelona, Spain

^132^ University Medicine Greifswald, Department of Psychiatry and Psychotherapy, Greifswald, Mecklenburg-Vorpommern,

Germany

^133^ German Centre for Neurodegenerative Diseases (DZNE), Partner Site Rostock/Greifswald, Greifswald, Mecklenburg-Vorpommern, Germany

^134^ University of Coimbra, Department of Psychiatry, Coimbra, Portugal

^135^ University College London, Division of Psychiatry, London, UK

^136^ Hospital de Clínicas de Porto Alegre, Laboratory of Developmental Psychiatry, Porto Alegre, RS, Brazil

^137^ Universidade Federal do Rio Grande do Sul, Department of Genetics, Porto Alegre, RS, Brazil

^138^ University of Melbourne, Department of Psychiatry, Melbourne Medical School, Melbourne, Australia

^139^ University of Münster, Department of Psychiatry, Münster, Germany

^140^ Assistance Publique - Hôpitaux de Paris, Department of Psychiatry and Addiction Medicine, Paris, France

^141^ FondaMental Foundation, Paris Bipolar and TRD Expert Centres, Paris, France

^142^ INSERM, UMR-S1144 Team 1 : Biomarkers of relapse and therapeutic response in addiction and mood disorders, Paris, France

^143^ Université Paris Cité, Psychiatry, Paris, France

^144^ University of Münster, Institute of Epidemiology and Social Medicine, Münster, Nordrhein-Westfalen, Germany

^145^ Mayo Clinic, Health Sciences Research, Rochester, MN, USA

^146^ State University of New York Downstate Medical Center, Department of Psychiatry and Behavioral Sciences, New York, NY, USA

^147^ Max Planck Institute of Psychiatry, Department of Translational Research in Psychiatry, Munich, Germany

^148^ University of Michigan, Center for Statistical Genetics and Department of Biostatistics, Ann Arbor, MI, USA

^149^ UMC Utrecht Brain Center, Psychiatry, Utrecht, Netherlands

^150^ Universitat Autònoma de Barcelona, Department of Psychiatry and Legal Medicine, Barcelona, Spain

^151^ University of California San Diego, Department of Psychiatry, La Jolla, CA, USA

^152^ University of New South Wales, School of Psychology, Sydney, NSW, Australia

^153^ University Hospital, LMU Munich, Institute of Psychiatric Phenomics and Genomics (IPPG), Munich, Germany

^154^ The University of Queensland, Child Health Research Centre, Brisbane, QLD, Australia

^155^ UMC Utrecht Hersencentrum Rudolf Magnus, Department of Psychiatry, Utrecht, Netherlands

^156^ University of Granada, Mental Health Unit, Department of Psychiatry, Faculty of Medicine, Granada University Hospital Complex, Granada, Spain

^157^ CNRS GDR 3557, Institut de Psychiatrie, Paris, France

^158^ GHU Paris Psychiatrie et Neurosciences, Department of Evaluation, Prevention and Therapeutic innovation, Paris, France

^159^ Université de Paris, Institute of Psychiatry and Neuroscience of Paris (IPNP), INSERM U1266, Team Pathophysiology of psychiatric diseases, Paris, France

^160^ Research Centre Jülich, Institute of Neuroscience and Medicine (INM-1), Jülich, Germany

^161^ University Hospital Basel, Institute of Medical Genetics and Pathology, Basel, Switzerland

^162^ University of Basel, Department of Biomedicine, Basel, Switzerland

^163^ University of Bonn, School of Medicine & University Hospital Bonn, Institute of Human Genetics, Bonn, Germany

^164^ Trinity College Dublin, Neuropsychiatric Genetics Research Group, Dept of Psychiatry and Trinity Translational Medicine Institute, Dublin, Ireland

^165^ Cardiff University, Medical Research Council Centre for Neuropsychiatric Genetics and Genomics, Division of Psychological Medicine and Clinical Neurosciences, Cardiff, UK

^166^ University of Southern California, Department of Translational Genomics, Pasadena, CA, USA

^167^ Oslo University Hospital, Department of Medical Genetics, Oslo, Norway

^168^ University of Bergen, NORMENT, KG Jebsen Centre for Psychosis Research, Department of Clinical Science, Bergen, Norway

^169^ Central Institute of Mental Health, Medical Faculty Mannheim, Heidelberg University, Department of Genetic Epidemiology in Psychiatry, Mannheim, Germany

^170^ University of Marburg, Centre for Human Genetics, Marburg, Germany

^171^ Mayo Clinic, Department of Psychiatry & Psychology, Rochester, MN, USA

^172^ NorthShore University HealthSystem, Department of Psychiatry and Behavioral Sciences, Evanston, IL, USA

^173^ University of Chicago, Department of Psychiatry and Behavioral Neuroscience, Chicago, IL, USA

^174^ Medical University of Vienna, Department of Psychiatry and Psychotherapy, Vienna, Austria

^175^ University of Munich, Department of Psychiatry, Munich, Germany

^176^ University of New South Wales, School of Psychiatry, Sydney, NSW, Australia

^177^ Hospital de Clínicas de Porto Alegre, ADHD Outpatient Program, Adult Division, Porto Alegre, RS, Brazil

^178^ Universidade Federal do Rio Grande do Sul, Department of Psychiatry, Porto Alegre, RS, Brazil

^179^ Alexandru Obregia Clinical Psychiatric Hospital, Biometric Psychiatric Genetics Research Unit, Bucharest, Romania

^180^ University of Granada, Department of Psychiatry, Faculty of Medicine and Biomedical Research Centre (CIBM), Granada, Spain

^181^ University Regional Hospital. Biomedicine Institute (IBIMA), Mental Health Department, Málaga, Spain

^182^ Kaiser Permanente Northern California, Psychiatry, San Francisco, CA, USA

^183^ Poznan University of Medical Sciences, Psychiatric Genetics, Department of Psychiatry, Poznan, Poland

^184^ Max Planck Institute of Psychiatry, Munich, Germany

^185^ University of Worcester, Department of Psychological Medicine, Worcester, UK

^186^ University of Gothenburg, Department of Psychiatry and Neuroscience, Gothenburg, Sweden

^187^ UMC Utrecht Brain Center Rudolf Magnus, Psychiatry, Utrecht, Netherlands

^188^ University of California San Diego, Institute for Genomic Medicine, La Jolla, CA, USA

^189^ University of Toronto, Department of Psychiatry, Toronto, ON, Canada

^190^ Harvard TH Chan School of Public Health, Department of Epidemiology, Boston, MA, USA

^191^ Massachusetts General Hospital, Department of Psychiatry, Boston, MA, USA

^192^ Center for Research in Environmental Epidemiology (CREAL), Barcelona, Spain

^193^ University of Gothenburg, Institute of Neuroscience and Physiology, Gothenburg, Sweden

^194^ North East London NHS Foundation Trust, Psychiatry, Ilford, UK

^195^ Univ Paris Est Créteil, INSERM, AP-HP, IMRB, Translational Neuropsychiatry, DMU IMPACT, FHU ADAPT, Fondation FondaMental, Créteil, France

^196^ INSERM, Paris, France

^197^ Université Paris Est, Faculté de Médecine, Créteil, France

^198^ Massachusetts General Hospital, Psychiatric and Neurodevelopmental Genetics Unit, Boston, MA, USA

^199^ Stanford University, Psychiatry & Behavioral Sciences, Stanford, CA, USA

^200^ Broad Institute of MIT and Harvard, Stanley Center for Psychiatric Research, Cambridge, MA, USA

^201^ Massachusetts General Hospital, Analytical and Translational Genetics Unit, Cambridge, MA, USA

^202^ M. Sklodowska-Curie Cancer Center and Institute of Oncology, Cancer Epidemiology and Prevention, Warsaw, Poland

^203^ Lindner Center of HOPE, Research Institute, Mason, OH, USA

^204^ Columbia University College of Physicians and Surgeons, Psychiatry, New York, NY, USA

^205^ Queensland University of Technology, School of Psychology and Counseling, Brisbane, QLD, Australia

^206^ The University of Queensland, Queensland Brain Institute, Brisbane, QLD, Australia

^207^ University of Oslo, Institute of Clinical Medicine, Division of Mental Health and Addiction, Oslo, Norway

^208^ Amsterdam UMC, Vrije Universiteit and GGZ inGeest, Department of Psychiatry, Amsterdam, Netherlands

^209^ University of Granada, Department of Nursing, Faculty of Health Sciences and Biomedical Research Centre (CIBM), Granada, Spain

^210^ Norwegian University of Science and Technology - NTNU, Mental Health, Faculty of Medicine and Health Sciences, Trondheim, Norway

^211^ St Olavs University Hospital, Psychiatry, Trondheim, Norway

^212^ Aarhus University, Centre for Integrated Register-based Research, Aarhus, Denmark

^213^ Munich Cluster for Systems Neurology (SyNergy), Munich, Germany

^214^ University of Liverpool, Liverpool, UK

^215^ University of Pittsburgh, Psychiatry and Human Genetics, Pittsburgh, PA, USA

^216^ Erasmus University Medical Center, Psychiatry, Rotterdam, Netherlands

^217^ Rutgers University, RWJMS,NJMS,UBHC, Pisctatway, NJ, USA

^218^ Rutgers University, RWJMS,NJMS, Pisctatway, NJ, USA

^219^ Amsterdam UMC, Vrije Universiteit, Department of Psychiatry and Amsterdam Neuroscience, Amsterdam, Netherlands

^220^ Johns Hopkins University School of Medicine, Psychiatry, Baltimore, MD, USA

^221^ BioMarin Pharmaceuticals, Genetics, London, UK

^222^ University of Oxford, St Edmund Hall, Oxford, UK

^223^ University of Oxford, Department of Psychiatry, Oxford, UK

^224^ University Hospital Frankfurt, Department of Psychiatry, Psychosomatic Medicine and Psychotherapy, Frankfurt, Germany

^225^ Central Institute of Mental Health, Medical Faculty Mannheim, Heidelberg University, Department of Genetic Epidemiology in Psychiatry, Mannheim, Baden-Württemberg, Germany

^226^ University of Granada, Department of Biochemistry and Molecular Biology II and Institute of Neurosciences, Biomedical Research Centre (CIBM), Granada, Spain

^227^ Harvard TH Chan School of Public Health, Department of Environmental Health, Boston, MA, USA

^228^ McGill University, Faculty of Medicine, Department of Neurology and Neurosurgery, Montreal, QC, Canada

^229^ Montreal Neurological Institute and Hospital, Montreal, QC, Canada

^230^ Instituto de Ciencias Biomedicas Universidade de Sao Paulo, Department of Physiology and Biophysics, São Paulo, SP, Brazil

^231^ Johns Hopkins University School of Medicine, Department of Psychiatry and Behavioral Sciences, Baltimore, MD, USA

^232^ National Institute of Mental Health, Human Genetics Branch, Intramural Research Program, Bethesda, MD, USA

^233^ University Medical Center Göttingen, Department of Psychiatry and Psychotherapy, Göttingen, Germany

^234^ University of Bologna, Department of Biomedical and NeuroMotor Sciences, Bologna, Italy

^235^ National Cancer Institute, Division of Cancer Epidemiology and Genetics, Bethesda, MD, USA

^236^ Kaiser Permanente Washington, Behavioral Health Services, Seattle, WA, USA

^237^ Icahn School of Medicine at Mount Sinai, Department of Neuroscience, New York, NY, USA

^238^ Massachusetts General Hospital, Psychiatric and Neurodevelopmental Genetics Unit (PNGU), Boston, MA, USA

^239^ King’s College London, Institute of Psychology, Psychiatry & Neuroscience, London, UK

^240^ Baylor College of Medicine, Houston, Menninger Department of Psychiatry and Behavioral Sciences, Houston, TX, USA

^241^ IRCCS Santa Lucia Foundation, Rome, Laboratory of Neuropsychiatry, Rome, Italy

^242^ Nofer Institute of Occupational Medicine, Department of Environmental Epidemiology, Lodz, Poland

^243^ University of California, San Diego, Center for Behavioral Genomics, Department of Psychiatry, La Jolla, CA, USA

^244^ McGill University, Department of Psychiatry, Montreal, QC, Canada

^245^ Centre for Addiction and Mental Health, Molecular Brain Science, Toronto, ON, Canada

^246^ University Medicine Greifswald, Institute for Community Medicine, Greifswald, Mecklenburg-Vorpommern, Germany

^247^ Columbia University College of Physicians and Surgeons, New York, NY, USA

^248^ New York State Psychiatric Institute, Division of Translational Epidemiology, New York, NY, USA

^249^ Stanford University, Department of Psychiatry and Behavioral Sciences, Stanford, CA, USA

^250^ University of Toronto, Institute of Medical Science, Toronto, ON, Canada

^251^ Centre for Addiction and Mental Health, Molecular Brain Science, Campbell Family Mental Health Research Institute, Toronto, ON, Canada

^252^ University of Toronto, Laboratory Medicine and Pathobiology, Toronto, ON, Canada

^253^ iPSYCH, The Lundbeck Foundation Initiative for Integrative Psychiatric Research, Aarhus, Denmark

^254^ Australian National University, Center of Mental Health Research, Canberra, Australia

^255^ Johns Hopkins Bloomberg School of Public Health, Department of Mental Health, Baltimore, MD, USA

^256^ Mental Health Centre Copenhagen, Danish Research Institute for Suicide Prevention, Copenhagen, Denmark

^257^ Broad Institute, Program in Medical and Population Genetics, Cambridge, MA, USA

^258^ University of Tartu, Estonian Genome Center, Institute of Genomics, Tartu, Estonia

^259^ SUNY Upstate Medical University, Department of Psychiatry and Behavioral Sciences, Syracuse, NY, USA

^260^ Statens Serum Institut, Center for Neonatal Screening, Department for Congenital Disorders, Copenhagen, Denmark

^261^ National Taiwan University Hospital and College of Medicine, Department of Psychiatry, Taipei, Taiwan

^262^ King’s College London, Department of Medical & Molecular Genetics, London, UK

^263^ Janssen Research & Development, LLC, Neuroscience, Titusville, NJ, USA

^264^ Aarhus University Hospital, Risskov, Psychosis Research Unit, Aarhus, Denmark

^265^ Copenhagen University Hospital, Mental Health Center Copenhagen, Copenhagen, Denmark

^266^ QIMR Berghofer Medical Research Institute, Department of Population Health, Brisbane, QLD, Australia

^267^ University of Edinburgh, Institute for Genetics and Molecular Medicine, Edinburgh, UK

^268^ University of Edinburgh, Centre for Clinical Brain Sciences, Edinburgh, UK

^269^ Regeneron Genetics Center, Analytical Genetics and Data Science, Tarrytown, NY, USA

^270^ University of California San Diego, Department of Psychiatry and School of Public Health, La Jolla, CA, USA

^271^ University of Copenhagen, Department of Clinical Medicine, Copenhagen, Denmark

^272^ University of Copenhagen, Lundbeck Foundation GeoGenetics Centre, GLOBE Institute,, Copenhagen, Denmark

^273^ University of Iowa, Department of Psychiatry, Iowa City, IA, USA

^274^ University of Utah School of Medicine, Biomedical Informatics, Salt Lake City, UT, USA

^275^ Durham Veterans Affairs Health Care System, VISN 6 Mid-Atlantic Mental Illness Research, Education, and Clinical Center, Durham, NC, USA

^276^ Duke University School of Medicine, Department of Psychiatry and Behavioral Sciences, Durham, NC, USA

^277^ Vanderbilt University Medical Center, Department of Biomedical Informatics, Nashville, TN, USA

^278^ Vanderbilt University Medical Center, Department of Psychiatry and Behavioral Sciences, Nashville, TN, USA

### MVP Suicide Exemplar Workgroup

Silvia Crivelli, Ph.D. (Lawrence Berkeley National Laboratory), Michelle F. Dennis, B.A. (Durham Veterans Affairs Health Care System & Duke University School of Medicine), Phillip D. Harvey, Ph.D. (University of Miami Miller School of Medicine, Miami, FL), Bruce W. Carter (VA Medical Center), Jennifer E. Huffman, Ph.D. (Massachusetts Veterans Epidemiology Research and Information Center, VA Boston Healthcare System), Daniel Jacobson, Ph.D. (Oak Ridge National Laboratory), Ravi Madduri, Ph.D. (Argonne National Laboratory), Maren K. Olsen, Ph.D. (Duke University School of Medicine), and John Pestian, Ph.D. (Oak Ridge National Laboratory).

### Veterans Administration Million Veteran Program (MVP)

J. Michael Gaziano, M.D., M.P.H. (co-chair, VA Boston Healthcare System), Sumitra Muralidhar, Ph.D. (co-chair, U.S. Department of Veterans Affairs), Rachel Ramoni, D.M.D., Sc.D. (U.S. Department of Veterans Affairs), Jean Beckham, Ph.D. (Durham VA Medical Center), Kyong-Mi Chang, M.D. (Philadelphia VA Medical Center), Christopher J. O’Donnell, M.D., M.P.H. (VA Boston Healthcare System), Philip S. Tsao, Ph.D. (VA Palo Alto Health Care System), James Breeling, M.D. (Ex-Officio, U.S. Department of Veterans Affairs), Grant Huang, Ph.D. (Ex-Officio, U.S. Department of Veterans Affairs), and J.P. Casas Romero, M.D., Ph.D. (Ex-Officio, VA Boston Healthcare System). MVP Program Office: Sumitra Muralidhar, Ph.D., and Jennifer Moser, Ph.D., both of U.S. Department of Veterans Affairs. MVP Recruitment/Enrollment: Recruitment/Enrollment Director/Deputy Director, Boston—Stacey B. Whitbourne, Ph.D., Jessica V. Brewer, M.P.H. (VA Boston Healthcare System). MVP Coordinating Centers: Clinical Epidemiology Research Center (CERC), West Haven—Mihaela Aslan, Ph.D. (West Haven VA Medical Center). Cooperative Studies Program Clinical Research Pharmacy Coordinating Center, Albuquerque—Todd Connor, Pharm.D., Dean P. Argyres, B.S., M.S. (New Mexico VA Health Care System). Genomics Coordinating Center, Palo Alto—Philip S. Tsao, Ph.D. (VA Palo Alto Health Care System). MVP Boston Coordinating Center, Boston—J. Michael Gaziano, M.D., M.P.H. (VA Boston Healthcare System). MVP Information Center, Canandaigua—Brady Stephens, M.S. (Canandaigua VA Medical Center). VA Central Biorepository, Boston—Mary T. Brophy, M.D., M.P.H., Donald E. Humphries, Ph.D., Luis E. Selva, Ph.D. (VA Boston Healthcare System). MVP Informatics, Boston—Nhan Do, M.D., Shahpoor Shayan (VA Boston Healthcare System). MVP Data Operations/Analytics, Boston—Kelly Cho, Ph.D. (VA Boston Healthcare System). MVP Science: Science Operations—Christopher J. O’Donnell, M.D., M.P.H. (VA Boston Healthcare System). Genomics Core— Christopher J. O’Donnell, M.D., M.P.H., Saiju Pyarajan, Ph.D. (VA Boston Healthcare System), Philip S. Tsao, Ph.D. (VA Palo Alto Health Care System). Phenomics Core—Kelly Cho, M.P.H., Ph.D. (VA Boston Healthcare System). Data and Computational Sciences—Saiju Pyarajan, Ph.D. (VA Boston Healthcare System). Statistical Genetics—Elizabeth Hauser, Ph.D. (Durham VA Medical Center). Yan Sun, Ph.D. (Atlanta VA Medical Center). Hongyu Zhao, Ph.D. (West Haven VA Medical Center. Current MVP Local Site Investigators: Peter Wilson, M.D. (Atlanta VA Medical Center); Rachel McArdle, Ph.D. (Bay Pines VA Healthcare System); Louis Dellitalia, M.D. (Birmingham VA Medical Center); Kristin Mattocks, Ph.D., M.P.H. (Central Western Massachusetts Healthcare System); John Harley, M.D., Ph.D. (Cincinnati VA Medical Center); Clement J. Zablocki (VA Medical Center); Jeffrey Whittle, M.D., M.P.H.; Frank Jacono, M.D. (VA Northeast Ohio Healthcare System); Jean Beckham, Ph.D. (Durham VA Medical Center); Edith Nourse Rogers Memorial Veterans Hospital; Salvador Gutierrez, M.D. (Edward Hines, Jr. VA Medical Center); Gretchen Gibson, D.D.S., M.P.H. (Veterans Health Care System of the Ozarks); Kimberly Hammer, Ph.D. (Fargo VA Health Care System); Laurence Kaminsky, Ph.D. (VA Health Care Upstate New York); Gerardo Villareal, M.D. (New Mexico VA Health Care System); Scott Kinlay, M.B.B.S., Ph.D. (VA Boston Healthcare System); Junzhe Xu, M.D. (VA Western New York Healthcare System); Mark Hamner, M.D. (Ralph H. Johnson VA Medical Center); Roy Mathew, M.D. (Columbia VA Health Care System); Sujata Bhushan, M.D. (VA North Texas Health Care System); Pran Iruvanti, DO, Ph.D. (Hampton VA Medical Center); Michael Godschalk, M.D. (Richmond VA Medical Center); Zuhair Ballas, M.D. (Iowa City VA Health Care System); Douglas Ivins, M.D. (Eastern Oklahoma VA Health Care System); Stephen Mastorides, M.D. (James A. Haley Veterans’ Hospital); Jonathan Moorman, M.D., Ph.D. (James H. Quillen VA Medical Center); Saib Gappy, M.D. (John D. Dingell VA Medical Center); Jon Klein, M.D., Ph.D. (Louisville VA Medical Center); Nora Ratcliffe, M.D. (Manchester VA Medical Center); Hermes Florez, M.D., Ph.D. (Miami VA Health Care System); Olaoluwa Okusaga, M.D. (Michael E. DeBakey VA Medical Center); Maureen Murdoch, M.D., M.P.H. (Minneapolis VA Health Care System); Peruvemba Sriram, M.D. (N FL/S GA Veterans Health System); Shing Shing Yeh, Ph.D., M.D. (Northport VA Medical Center); Neeraj Tandon, M.D. (Overton Brooks VA Medical Center); Darshana Jhala, M.D. (Philadelphia VA Medical Center); Samuel Aguayo, M.D. (Phoenix VA Health Care System); David Cohen, M.D. (Portland VA Medical Center); Satish Sharma, M.D. (Providence VA Medical Center); Suthat Liangpunsakul, M.D., M.P.H. (Richard Roudebush VA Medical Center); Kris Ann Oursler, M.D. (Salem VA Medical Center); Mary Whooley, M.D. (San Francisco VA Health Care System); Sunil Ahuja, M.D. (South Texas Veterans Health Care System); Joseph Constans, Ph.D. (Southeast Louisiana Veterans Health Care System); Paul Meyer, M.D., Ph.D. (Southern Arizona VA Health Care System); Jennifer Greco, M.D. (Sioux Falls VA Health Care System); Michael Rauchman, M.D. (St. Louis VA Health Care System); Richard Servatius, Ph.D. (Syracuse VA Medical Center); Melinda Gaddy, Ph.D. (VA Eastern Kansas Health Care System); Agnes Wallbom, M.D., M.S. (VA Greater Los Angeles Health Care System); Timothy Morgan, M.D. (VA Long Beach Healthcare System); Todd Stapley, D.O. (VA Maine Healthcare System); Scott Sherman, M.D., M.P.H. (VA New York Harbor Healthcare System); George Ross, M.D. (VA Pacific Islands Health Care System); Philip Tsao, Ph.D. (VA Palo Alto Health Care System); Patrick Strollo Jr., M.D. (VA Pittsburgh Health Care System); Edward Boyko, M.D. (VA Puget Sound Health Care System); Laurence Meyer, M.D., Ph.D. (VA Salt Lake City Health Care System); Samir Gupta, M.D., M.S.C.S. (VA San Diego Healthcare System); Mostaqul Huq, Pharm.D., Ph.D. (VA Sierra Nevada Health Care System); Joseph Fayad, M.D. (VA Southern Nevada Healthcare System); Adriana Hung, M.D., M.P.H. (VA Tennessee Valley Healthcare System); Jack Lichy, M.D., Ph.D. (Washington, DC VA Medical Center); Robin Hurley, M.D. (W.G., Bill Hefner VA Medical Center); Brooks Robey, M.D. (White River Junction VA Medical Center); and Robert Striker, M.D., Ph.D. (William S. Middleton Memorial Veterans Hospital).

